# Global neuropathologic severity of Alzheimer’s disease and locus coeruleus vulnerability influences plasma phosphorylated tau levels

**DOI:** 10.1101/2022.10.12.22280971

**Authors:** Melissa E. Murray, Christina M. Moloney, Naomi Kouri, Jeremy A. Syrjanen, Billie J. Matchett, Darren M. Rothberg, Jessica F. Tranovich, Tiffany N. Hicks Sirmans, Heather J. Wiste, Baayla D. C. Boon, Aivi T. Nguyen, R. Ross Reichard, Dennis W. Dickson, Val J. Lowe, Jeffrey L Dage, Ronald C. Petersen, Clifford R. Jack, David S. Knopman, Prashanthi Vemuri, Jonathan Graff-Radford, Michelle M. Mielke

**Author notes:** Correspondence should be addressed to: Melissa E. Murray, PhD., Associate Professor of Neuroscience, Translational Neuropathology Laboratory, Mayo Clinic, 4500 San Pablo Road, Jacksonville FL 32224, Michelle M. Mielke, Ph.D., Chair, Department of Epidemiology and Prevention, Division of Public Health Sciences, Wake Forest University School of Medicine, 525 Vine, 5th floor, Winston-Salem, NC 27157.

## Abstract

Advances in ultrasensitive detection of phosphorylated tau (p-tau) in plasma has enabled the use of blood tests to measure Alzheimer’s disease (AD) biomarker changes. Examination of postmortem brains of participants with antemortem plasma p-tau levels remains critical to understanding comorbid and AD-specific contribution to these biomarker changes. We analyzed 35 population-based Mayo Clinic Study of Aging participants with plasma p-tau at threonine181 and threonine217 (p-tau181, p-tau217) available within 3 years of death. Autopsied participants included cognitively unimpaired, mild cognitive impairment, AD dementia, and non-AD neurodegenerative disorders. Global neuropathologic scales of tau, amyloid-β, TDP-43, and cerebrovascular disease were examined. Regional digital pathology measures of tau (phosphorylated threonine181 and 217 [pT181, pT217]) and amyloid-β (6F/3D) were quantified in hippocampus and parietal cortex. Neurotransmitter hubs reported to influence development of tangles (nucleus basalis of Meynert) and amyloid-β plaques (locus coeruleus) were evaluated. The strongest regional associations were with parietal cortex for tau burden (p-tau181 R=0.55, p=0.003; p-tau217 R=0.66, p<0.001) and amyloid-β burden (p-tau181 R=0.59, p<0.001; p-tau217 R=0.71, p<0.001). Linear regression analysis of global neuropathologic scales explained 31% of variability in plasma p-tau181 (R^2^=0.31) and 59% in plasma p-tau217 (R^2^=0.59). Neither TDP-43 nor cerebrovascular disease global scales independently contributed to variability. Global scales of tau pathology (β-coefficient=0.060, p=0.016) and amyloid-β pathology (β-coefficient=0.080, p<0.001) independently predicted plasma p-tau217 when modeled together with co-pathologies, but only amyloid-β (β-coefficient=0.33, p=0.021) significantly predicted plasma p-tau181. While nucleus basalis of Meynert neuron count/mm^2^ was not associated with plasma p-tau levels, a lower locus coeruleus neuron count/mm^2^ was associated with higher plasma p-tau181 (R=-0.50, p=0.007) and higher plasma p-tau217 (R=-0.55, p=0.002). Cognitive scores (R^2^=0.31-0.34) were predicted by the global tau scale, but not by the global amyloid-β scale or plasma p-tau when modeled simultaneously. Higher soluble plasma p-tau levels may be the result of an intersection between insoluble deposits of amyloid-β and tau accumulation in brain and may be associated with locus coeruleus degeneration.

## Background

Recent advances in technology have enabled ultrasensitive detection of plasma-derived phosphorylated-tau (p-tau) levels as a potential minimally-invasive Alzheimer’s disease (AD) biomarker [1-4]. Plasma p-tau levels could provide a more feasible AD biomarker than neuroimaging or lumbar puncture at the population level for diagnosis or screening purposes [2]. Postmortem validation of antemortem plasma p-tau changes is critical for understanding the strength of the relationship between neuropathology and plasma p-tau levels, and thus, translation to the clinic [2, 5]. Autopsy series have demonstrated higher plasma p-tau levels in patients with AD dementia, compared to non-AD dementias, and high accuracy for predicting AD dementia [6-8]. Interestingly, non-disease controls and non-AD cases were shown to not differ in plasma p-tau levels, suggesting an AD-specific biomarker increase even when comparing to other tauopathies [1, 9, 10].

Comparison of plasma p-tau at threonine181 (p-tau181) and threonine217 (p-tau217) levels with brain-derived global tau scales provides important insight into neuropathologic temporality and individual variability [4, 11]. Neuroimaging studies suggest plasma p-tau levels reliably predict in vivo positron emission tomography (PET) assessments of both tau and amyloid-β [1, 8], but the correlation with amyloid-β may be stronger – especially in individuals with elevated amyloid-β PET measures (i.e. amyloid-PET positive) [3]. The stronger relationship in amyloid-PET positive individuals could be an inferred reflection of greater tau pathology in brain or an amyloid-β-specific neuronal reaction influencing soluble p-tau production [12, 13]. Thus, to inform clinical interpretation we investigated the contribution of tau and amyloid-β neuropathology to plasma p-tau181 and p-tau217 levels and identified whether other sources of variability contribute to plasma p-tau levels. As plasma p-tau levels are derived from circulating blood, we hypothesized that global scales of neuropathology would demonstrate a stronger association with plasma p-tau levels compared to regional measures digitally quantified from the hippocampus or parietal cortex. We further assessed the impact of medical comorbidities [14] and common co-existing pathologies in the aging brain, including cerebrovascular disease [15] and TAR DNA binding protein 43 (TDP-43) [16], on plasma p-tau levels. Our overall goal was to examine clinicopathologic contributors to plasma p-tau181 and p-tau217 levels using global scales and regional measures to uncover sources of variability to help inform interpretation in the context of aging and neurodegeneration.

## Materials and Methods

### Participants

The Mayo Clinic Study of Aging (MCSA) is a population based, prospective study of residents living in Olmsted County, Minnesota. MCSA participants aged 70-89 were enumerated from the Rochester Epidemiology Project (REP) medical records-linkage system in 2004 and recruitment was extended in 2012 to participants aged 50 and older [17]. MCSA visits included an interview by a study coordinator, physician examination, cognitive testing, and a blood draw completed on the same day. The inclusion criteria for this study were MCSA participants who had undergone autopsy and who had plasma p-tau181 and p-tau217 levels within 3 years of death. To assess the relationship between neuropathology and cognition, Clinical Dementia Rating (CDR) [18] and Mini-Mental State Examination (MMSE) [19] were evaluated if testing occurred within 3 years of death.

### Procedures

Blood was collected in-clinic after an overnight fast. The blood was centrifuged, resulting plasma aliquoted, and stored at -80°C. Both p-tau181 and p-tau217 levels were measured in duplicate on a streptavidin small spot plate using the meso scale discovery (MSD) platform by electrochemiluminescence using proprietary assays developed by Lilly Research Laboratories, as previously described [20]. Levels of creatinine, alanine transaminase (ALT), and aspartate aminotransferase (AST) were abstracted from the medical records at the time closest to the MCSA visit with the blood draw for the p-tau levels. Only serum creatinine (mg/dL), ALT (U/L), and AST (U/L) levels identified within 3 years of the plasma p-tau blood draw were abstracted.

Neuropathologic sampling followed Consortium to Establish a Registry for Alzheimer’s Disease (CERAD) recommendations and National Institute on Aging-Alzheimer’s Association criteria for AD neuropathologic change assessment [21, 22]. Formalin-fixed, paraffin-embedded 5-µm-thick tissue sections were stained with hematoxylin and eosin, as well as Bielschowsky silver stain. Immunohistochemistry was performed on a Thermo Fisher Lab Vision 480S autostainer with 3,3-diaminobenzidine as chromogen. Antibodies against tau, amyloid-β, α-synuclein, and TDP-43 were used for neuropathologic evaluation (**Table S1**) [22-24]. Cases were also evaluated for global scales that included Braak stage, Thal phase, limbic-predominant age-related TDP-43 encephalopathy neuropathologic change (LATE-NC), and Kalaria cerebrovascular disease score [15, 16, 25, 26]. Diffuse plaques and neuritic plaques were evaluated using a 4-point semi-quantitative scale: none, mild, moderate, and severe. Neuropathologic diagnoses were rendered by two board-certified neuropathologists (RRR and ATN). Neuropathologic grouping for graphical visualization was prioritized as follows: progressive supranuclear palsy (n=2) [27], AD (n=9) [22], argyrophilic grain disease (n=3) [28], primary age-related tauopathy (n=14) [29], and pathological aging (n=7) [30, 31] (**Fig. S1**). Participants were assigned the neuropathologic diagnosis of AD if they had a Braak stage ≥IV and had at least moderate neuritic plaques. Primary age-related tauopathy was assigned if they had a Braak stage ≤IV and Thal phase ≤2. As part of ongoing efforts to investigate AD biomarkers [32, 33], we next assign pathological aging if they had a Braak ≤III and at least moderate diffuse plaques with no more than moderate neuritic plaques. During evaluation of antemortem contributors to plasma p-tau variability, a single outlier argyrophilic grain disease case was identified that was excluded from subsequent correlation and modeling analyses of neuropathology and cognition.

To clarify nomenclature when describing phosphorylated tau, “p-tau” is used for biomarker levels and “pT” for neuropathologic burden. The terms neuropathology and pathology are exclusively used when referencing examination of brain tissue and not used in reference to plasma p-tau biomarker changes. Digital pathology was used to quantify histopathologic burden of tau (pT181, pT217) and amyloid-β (6F/3D) in inferior parietal cortex and CA1-subiculum of hippocampus, as described in **Supplementary Methods** (**Tables S2-S3**). The parietal cortex was chosen as a region affected by accumulation of tau pathology in advanced AD (Braak stage >IV), but with limited involvement hypothesized to be influenced by age-related tau pathology [29]. The CA1 and subiculum subsectors of the hippocampus were specifically chosen to enhance relevance to AD, as they are highly vulnerable to tau pathology in AD compared to CA2 involvement more readily observed in primary tauopathies and age-related tauopathies [34, 35]. Neurotransmitter hubs reported to influence development of neurofibrillary tangles (i.e., nucleus basalis of Meynert) and amyloid-β plaques (i.e., locus coeruleus) were additionally evaluated for neuronal count/mm^2^ on hematoxylin and eosin using pattern recognition digital pathology software, as described in in **Supplementary Methods** (**Table S4, Fig. S2-S4**).

### Statistical Analysis

The statistical analysis consisted of five parts: a descriptive summary table giving median (25^th^, 75^th^ percentiles) for the variables included in our study, receiver operating characteristic (ROC) curves with corresponding area under the curves (AUC), scatter plots, Spearman correlations, and linear regression models. The ROC/AUC analysis was run to ascertain the ability of continuous plasma p-tau181 and p-tau217 levels to predict intermediate-to-high from none-to-low AD neuropathologic change. To perform this analysis, logistic regression models using the plasma markers and cognitive scores were individually run, and the predicted values from these models were used to create the ROC curves and compute the AUCs. We also computed confidence intervals for the AUCs using bootstrap resampling with 10,000 replicates. The latter three analysis methods were used to investigate the relationship between the various measures of neuropathology and plasma p-tau181 and p-tau217 as well as the relationship of other important factors (i.e., age, creatinine, etc.) with these plasma markers. In the linear models, the plasma markers served as the outcomes and β-coefficients, confidence intervals, and p-values were computed. All variables are shown for each model. Neuropathology variables were used as predictors and adjusted-*R*^2^ values were computed. Global neuropathology variables used as predictors included Braak stage (tau), Thal phase (amyloid-β), LATE-NC (TDP-43), and Kalaria cerebrovascular disease score. Global scales of tau and amyloid-β provide information on the topographic distribution of pathology (i.e., presence), but may not reflect severity of pathology. To quantitatively assess tau and amyloid-β pathology in corticolimbic regions, digital pathology was employed to enable a wider range of severity to be measured. Thus, regional neuropathology variables used as predictors included pT181, pT217, and 6F3D quantitatively measured in CA1-subiculum of the hippocampus and parietal cortex. The standard cutoff of p<0.05 was used to determine statistical significance. All statistical analyses were performed with R version 3.6.2 (R Foundation for Statistical Computing, Vienna, Austria) and SAS version 9.4 (SAS Institute, Cary, NC).

## Results

Among the 35 autopsy cases, the median (interquartile range) age of death was 86 (81, 89) years, 20 were male (57%) and all self-identified as non-Hispanic white (**Table 1**). The median time from plasma draw to death was 2.0 (1.4, 2.3) years.

**Table 1.** Participant characteristics summary table by neuropathologic grouping.

|  | **PSP (n=2)** | **AD (n=9)** | **AGD (n=2\*)** | **PART (n=14)** | **PA (n=7)** |
| --- | --- | --- | --- | --- | --- |
| Age range at plasma, years | 75-84 | 80-94 | 85-94 | 45-99 | 65-89 |
| Age range at death, years | 75-84 | 80-99 | 85-94 | 50-99 | 65-89 |
| Postmortem interval, hours | 4 (4,5) | 13 (9,18) | 20 (19,21) | 20 (18,22) | 14 (11,21) |
| Females | 0 (0%) | 4 (44%) | 1 (50%) | 8 (57%) | 2 (29%) |
| APOE ε4 positivity | 0 (0%) | 4 (44%) | 1 (50%) | 2 (14%) | 3 (43%) |
| Clinical Dementia Rating, 0-18 | 0.5 (0,1) | 4 (0.5,7) | 2 (1,3) | 0 (0,0) | 0 (0,0.5) |
| Mini-Mental State Examination, 0-30 | 29 (28,30) | 23 (23,27) | 28 (27,28) | 28 (28,29) | 28 (28,30) |
| Tau-PET parietal SUVR | n/a | 1.2 (1.1,1.3) | n/a | 1.0 (1.0,1.1) | 1.1 (1.1,1.1) |
| **Fluid levels** |  |  |  |  |  |
| MSD plasma p-tau181, pg/mL | 1.6 (1.3,1.9) | 3.4 (2.5,4.3) | 1.7 (1.6,1.9) | 1.2 (0.9,1.5) | 1.3 (1.1,1.8) |
| MSD plasma p-tau217, pg/mL | 0.25 (0.21,0.29) | 0.69 (0.52,0.75) | 0.18 (0.17,0.20) | 0.15 (0.12,0.20) | 0.19 (0.17,0.29) |
| Creatinine, mg/dL | 1.0 (1.0,1.0) | 1.2 (0.9,1.3) | 1.4 (1.2,1.6) | 1.0 (1.0,1.4) | 1.1 (1.1,1.3) |
| ALT level, U/L | n/a | 22 (15,24) | 21 (21,21) | 21 (18,27) | 19 (18,47) |
| AST level, U/L | 24 (21,27) | 23 (19,29) | 18 (15,20) | 26 (23,31) | 28 (22,49) |
| **Global scales** |  |  |  |  |  |
| Braak stage, 0-VI | III (III,III) | IV (IV,V) | II (II,II) | II (II,III) | II (II,II) |
| 0 \| I \| II \| III \| IV \| V \| VI, counts | 0\|0\|0\|2\|0\|0\|0 | 0\|0\|0\|0\|5\|3\|1 | 0\|0\|2\|0\|0\|0\|0 | 1\|3\|4\|5\|1\|0\|0 | 0\|2\|4\|1\|0\|0\|0 |
| Thal phase, 0-5 | 3 (3,3) | 5 (4,5) | 0.5 (0,1) | 1 (1,2) | 3 (3,4) |
| 0 \| 1 \| 2 \| 3 \| 4 \| 5, counts | 0\|0\|0\|2\|0\|0 | 0\|0\|0\|2\|2\|5 | 1\|1\|0\|0\|0\|0 | 3\|6\|5\|0\|0\|0 | 0\|0\|0\|4\|3\|0 |
| Diffuse plaques, Moderate-Frequent | 2 (100%) | 9 (100%) | 1 (50%) | 3 (21%) | 7 (100%) |
| Neuritic plaques, Moderate-Frequent | 2 (100%) | 9 (100%) | 0 (0%) | 1 (7%) | 4 (57%) |
| LATE-NC stage, 0-3 | 0 (0,0) | 1 (0,2) | 1 (1,2) | 0 (0,0) | 0 (0,0) |
| Kalaria CVD score, 0-10 | 3 (3,3) | 4 (3,5) | 6 (5,6) | 2 (2,4) | 3 (2,5) |
| **Regional measures** |  |  |  |  |  |
| Parietal pT181, % | 0.27 (0.27,0.27) | 0.64 (0.54,0.93) | 0.27 (0.22,0.32) | 0.17 (0.12,0.22) | 0.11 (0.080,0.23) |
| Parietal pT217, % | 0.46 (0.38,0.55) | 0.63 (0.38,4.9) | 0.22 (0.20,0.25) | 0.080 (0.060,0.090) | 0.11 (0.10,0.12) |
| Parietal amyloid-β, % | 2.7 (2.5,3.0) | 2.7 (2.0,4.6) | 0.24 (0.16,0.32) | 0.16 (0.10,0.46) | 1.1 (0.43,1.5) |
| Hippocampal pT181, % | 0.58 (0.41,0.74) | 2.5 (1.9,3.9) | 0.84 (0.51,1.2) | 0.28 (0.18,0.34) | 0.34 (0.16,0.38) |
| Hippocampal pT217, % | 4.8 (3.0,6.6) | 21 (10,28) | 6.6 (3.5,9.7) | 2.3 (0.34,3.7) | 0.95 (0.67,2.7) |
| Hippocampal amyloid-β, % | 0.36 (0.25,0.47) | 0.64 (0.16,1.8) | 0.050 (0.040,0.050) | 0.050 (0.040,0.12) | 0.11 (0.060,0.34) |
| LC neurons/mm<sup>2</sup> | 25 (18,33) | 30 (22,33) | 22 (22,22) | 44 (38,52) | 45 (39,52) |
| nbM neurons/mm<sup>2</sup> | 14 (11,17) | 19 (14,19) | 30 (26,35) | 22 (17,27) | 20 (15,29) |

Data presented are median and interquartile range (25<sup>th</sup>, 75<sup>th</sup>). Age is presented in ranges to protect identity of participants. \*AGD case with high creatinine was not included. Acronyms: AD=Alzheimer's disease. AGD=Argyrophilic grains disease. ALT=alanine transaminase. AST=aspartate aminotransferase. CVD=cerebrovascular disease. CAA=cerebral amyloid angiopathy. LATE-NC=limbic-predominant age-related TDP-43 encephalopathy neuropathologic change. LC=locus coeruleus. MSD=meso scale discovery. p-tau=phosphorylated tau in plasma. n/a=not available. nbM=nucleus basalis of Meynert. PA=pathological aging. PART=primary age-related tauopathy. PSP=progressive supranuclear palsy. pT=phosphorylated threonine for immunohistochemical measures of tau.

**Table 2.**
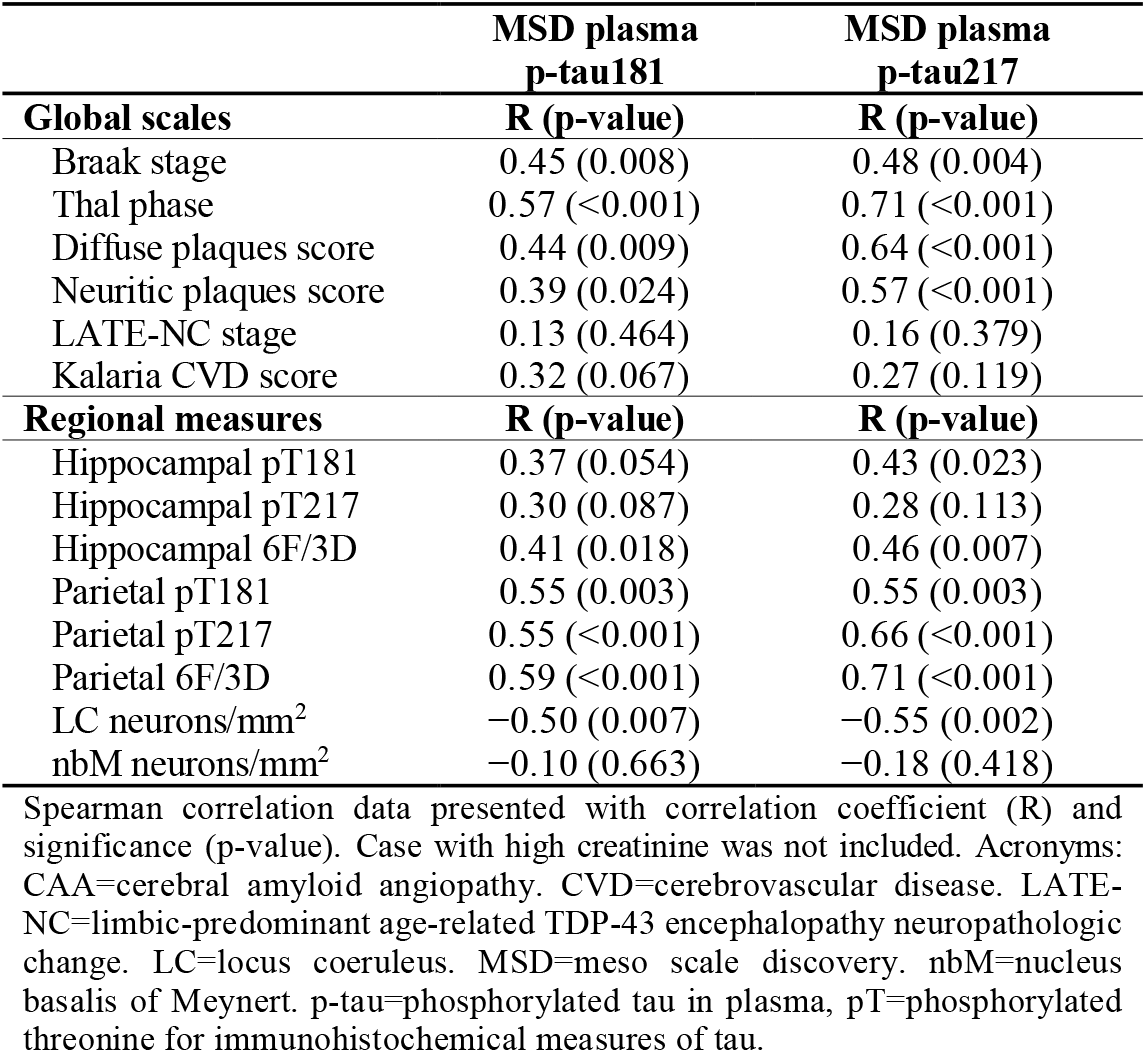
Evaluation of the relationship between global scales and regional neuropathologic measures with plasma p-tau levels.

|  | MSD plasma<br>p-tau181 | MSD plasma<br>p-tau217 |
| --- | --- | --- |
| Global scales | R (p-value) | R (p-value) |
| Braak stage | 0.45 (0.008) | 0.48 (0.004) |
| Thal phase | 0.57 (<0.001) | 0.71 (<0.001) |
| Diffuse plaques score | 0.44 (0.009) | 0.64 (<0.001) |
| Neuritic plaques score | 0.39 (0.024) | 0.57 (<0.001) |
| LATE-NC stage | 0.13 (0.464) | 0.16 (0.379) |
| Kalaria CVD score | 0.32 (0.067) | 0.27 (0.119) |
| Regional measures | R (p-value) | R (p-value) |
| Hippocampal pT181 | 0.37 (0.054) | 0.43 (0.023) |
| Hippocampal pT217 | 0.30 (0.087) | 0.28 (0.113) |
| Hippocampal 6F/3D | 0.41 (0.018) | 0.46 (0.007) |
| Parietal pT181 | 0.55 (0.003) | 0.55 (0.003) |
| Parietal pT217 | 0.55 (<0.001) | 0.66 (<0.001) |
| Parietal 6F/3D | 0.59 (<0.001) | 0.71 (<0.001) |
| LC neurons/mm<sup>2</sup> | -0.50 (0.007) | -0.55 (0.002) |
| nbM neurons/mm<sup>2</sup> | -0.10 (0.663) | -0.18 (0.418) |

Spearman correlation data presented with correlation coefficient (R) and significance (p-value). Case with high creatinine was not included. Acronyms: CAA=cerebral amyloid angiopathy. CVD=cerebrovascular disease. LATE-NC=limbic-predominant age-related TDP-43 encephalopathy neuropathologic change. LC=locus coeruleus. MSD=meso scale discovery. nbM=nucleus basalis of Meynert. p-tau=phosphorylated tau in plasma, pT=phosphorylated threonine for immunohistochemical measures of tau.

### Antemortem contributors evaluated for contribution to plasma p-tau variability

As previous studies suggested that comorbidities, including both kidney and liver disease, may affect p-tau biomarker levels [14, 36], we first examined Spearman correlations of creatinine, AST, and ALT with plasma p-tau levels (**Fig. S5**). One individual was identified as an outlier with high serum creatinine (3.7 mg/dL) and the highest plasma p-tau levels (p-tau181=10 pg/mL, p-tau217=1.3 pg/mL). This individual remained part of the initial analyses evaluating antemortem variability of creatinine, AST, ALT, age at plasma p-tau draw, and time from blood draw to death. However, this individual was removed from subsequent analyses investigating neuropathology and cognition. AST and ALT were not found to significantly associate with plasma p-tau levels, which is further described in **Supplemental Results**. We next examined the relationship between age at plasma p-tau draw and time from plasma draw to death with plasma p-tau levels (**Fig. S5**). Age at plasma draw did not correlate with plasma p-tau181 (R=0.25, p=0.142) or with plasma p-tau217 (R=0.27, p=0.121). Similarly, time from plasma to death did not correlate with plasma p-tau181 (R=-0.012, p=0.947) or with plasma p-tau217 (R=-0.027, p=0.879).

### Qualitative global scales of neuropathology evaluated as predictors of plasma p-tau

We next assessed the relationship between global scales of AD neuropathologic change and plasma p-tau levels (**Table 2**). A discernible threshold of increase visibly appeared to be between Braak stage III and stage IV (**Fig. 1A, C**), corresponding with the observation of cortical tau pathology by Braak stage IV [25, 37]. Braak stage associated with both plasma p-tau181 (R=0.45, p=0.008) and plasma p-tau217 (R=0.48, p=0.004). Visual inspection of Thal phase graphs suggested a biological effect of increase between Thal phase 3 and phase 4 (**Fig. 1B, D**), corresponding with observations of amyloid-β plaque pathology in brainstem by Thal phase 4 [26]. While Thal phase strongly associated with plasma p-tau181 (R=0.57, p<0.001), the correlation was even stronger with plasma p-tau217 (R=0.71, p<0.001). Similarly, diffuse plaque scores (p-tau181 R=0.44, p=0.009; p-tau217 R=0.64, p<0.001) and neuritic plaque scores (p-tau181 R=0.39, p=0.024; p-tau217 R=0.57, p<0.001) was more strongly associated with plasma p-tau217 compared to p-tau181 (**Table 2**).

**Figure 1.**
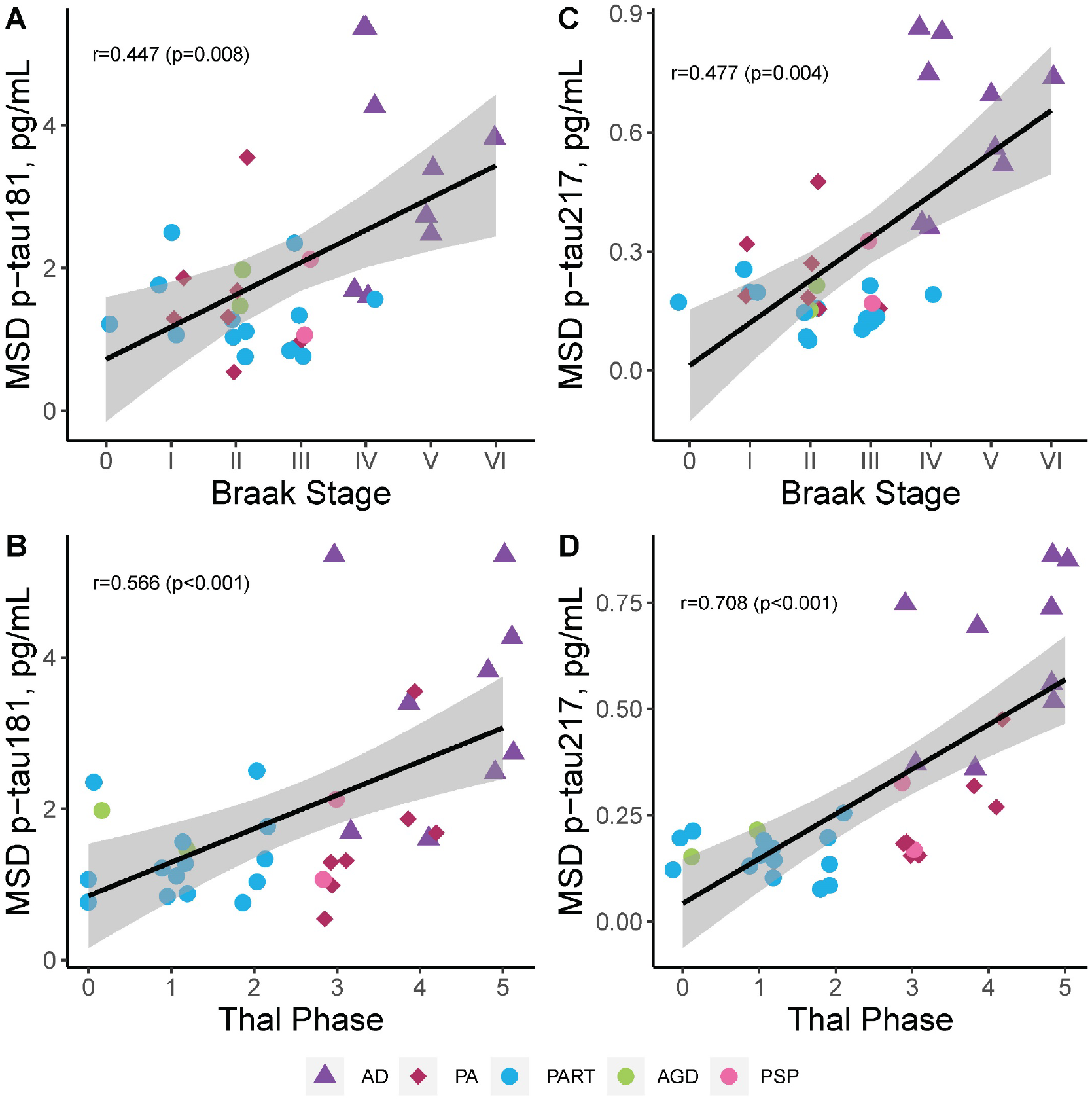
Neuropathologic evaluation of global tau and amyloid-β scales with plasma p-tau levels. We observed a strong association between the global tau scale (Braak stage [25]) and both p-tau181 (**A**) and plasma p-tau217 **(C)** across all cases studied. Visual inspection of graphs suggests a biological effect in plasma p-tau elevation between Braak stages III-IV **(A, C)**. We observed an even stronger relationship between the global amyloid scale (Thal phase [26]) and p-tau181 **(B)**, with the strongest association observed between Thal phase and plasma p-tau217 (D). Visual inspection of graphs suggests a biological effect in plasma p-tau elevation between Thal phase 3-4 **(B, D)**. P-tau181 **(A-B)** and p-tau217 **(C-D)** were examined across all individuals studied with AD shown as triangles, PA as diamonds, and primary tauopathies as circles. Spearman correlation and corresponding significance displayed. Case with high creatinine was not included. Trendline with 95% confidence interval was computed from a linear model. Data presented are Spearman correlation and corresponding significance. Acronyms: AD=Alzheimer’s disease. AGD=argyrophilic grain disease. MSD=meso scale discovery. PA=pathological aging. PART=primary age-related tauopathy. pg/mL=picograms per milliliter. PSP=progressive supranuclear palsy. p-tau=phosphorylated tau for plasma levels.

Based upon the surprising past [3, 38, 39] and current (**Table 2)** observations of a stronger association between amyloid-β in the brain and plasma p-tau, we sought to investigate the predictive relationship between neuropathologic variables as predictors and plasma p-tau levels as outcome. In consideration of the common observation of co-existing neuropathologies in the aging brain [15, 16, 40, 41], we included TDP-43 (LATE-NC [16]) and cerebrovascular disease (Kalaria score [15, 42]) in the regression model. Models were restricted to four predictors based upon sample size, but sensitivity analyses including time from plasma to death confirmed observations presented in **Table S5**. When examining global scales of neuropathology, 31% of the variability in p-tau181 was explained (R^2^=0.31). Thal phase (β-coefficient=0.33 [0.053, 0.60], p=0.021) was the main predictor of p-tau181. Neither Braak stage, LATE-NC stage, nor Kalaria cerebrovascular disease score contributed independently to p-tau181 levels. In contrast to p-tau181, global scales accounted for 59% of the variability in p-tau217 (R^2^=0.59). Both Thal phase (β-coefficient=0.080 [0.042, 0.12], p<0.001) and Braak stage (β-coefficient=0.060 [0.012, 0.11], p=0.016) independently predicted p-tau217 levels, but LATE-NC stage or Kalaria cerebrovascular disease score did not.

### Quantitative digital pathology measures evaluated as predictors of plasma p-tau

Multivariable regression analyses utilizing global scales of neuropathology provided further evidence that accumulating insoluble amyloid-β plaque pathology predicts higher soluble p-tau levels in plasma (**Table S5**). To further evaluate this perplexing relationship, quantitative regional measures of immunohistochemical burden of tau and amyloid-β using digital pathology were next evaluated in the CA1-subiculum hippocampal subsectors and parietal cortex (**Table 2**). The association of hippocampal burden with plasma p-tau181 was similar for tau pathology (pT181 R=0.37, p=0.054) and amyloid-β pathology (6F/3D R=0.40, p=0.018) (**Fig. S6**). However, the association of hippocampal burden with plasma p-tau217 was lower for tau pathology (pT217 R=0.28, p=0.113) than amyloid-β pathology (6F/3D R=0.46, p=0.007). The strength of the association of parietal cortex burden with plasma p-tau181 was robustly observed for both tau pathology (pT181 R=0.55, p=0.003; **Fig. 2A**) and amyloid-β pathology (6F/3D R=0.59, p<0.001; **Fig. 2B**). The strongest association observed between parietal cortex burden and plasma p-tau217 was for tau pathology (pT217 R=0.66, p<0.001; **Fig. 2C**) and amyloid-β pathology (6F/3D R=0.71, p<0.001; **Fig. 2D**).

**Figure 2.**
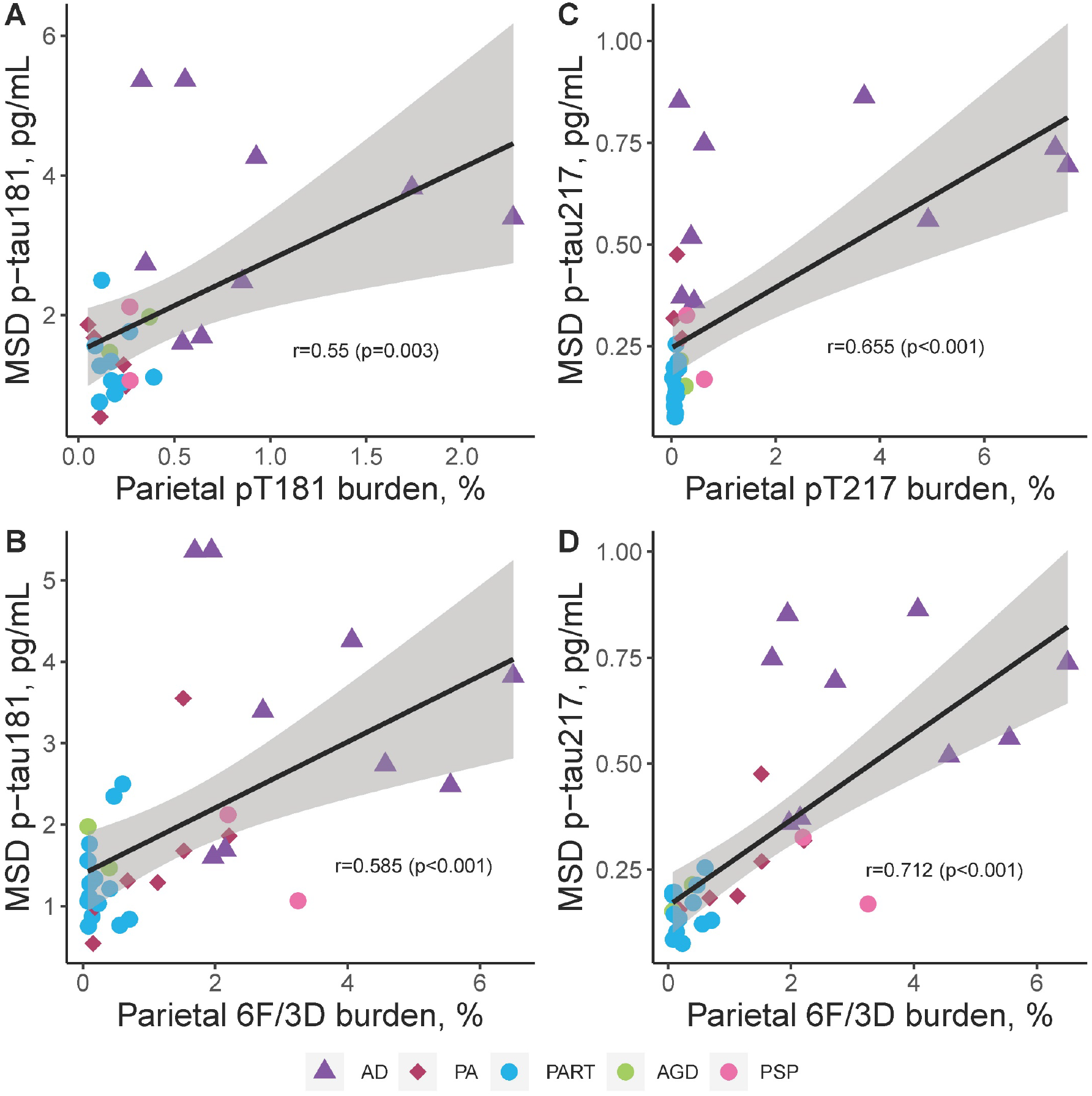
Neuropathologic evaluation of regional digital pathology measures of tau and amyloid-β pathology in comparison to plasma p-tau181 and p-tau217 in parietal cortex. Utilizing the same epitope to immunohistochemically evaluate regional tau pathology in inferior parietal cortex, tau burden measures were compared to p-tau plasma levels **(A, C)**. pT181 tau burden measures strongly associated with p-tau181 plasma levels **(A)**, with the association even stronger between pT217 and p-tau217 plasma levels **(C)**. Digital pathology measures of amyloid-β (6F/3D) were additionally compared to plasma p-tau levels **(B, D)**. Amyloid-β burden strongly associated with ptau-181 **(B)**. The strongest overall association of digital pathology measures was observed between amyloid-β (6F/3D) and p-tau217 **(D)**. P-tau181 **(A-B)** and p-tau217 **(C-D)** were examined across all individuals studied with AD shown as triangles, PA as diamonds, and primary tauopathies as circles. Spearman correlation and corresponding significance displayed. Case with high creatinine was not included. Trendline with 95% confidence interval was computed from a linear model. Data presented are Spearman correlation and corresponding significance. Acronyms: AD=Alzheimer’s disease. AGD=argyrophilic grain disease. MSD=meso scale discovery. PA=pathological aging. PART=primary age-related tauopathy. PSP=progressive supranuclear palsy. p-tau=phosphorylated tau for plasma levels. pT=phosphorylated threonine for immunohistochemical measures of tau.

To focus our investigation, we next performed linear modeling utilizing digital pathology burden measures derived from parietal cortex because this region was more strongly correlated with plasma p-tau levels compared to hippocampus (**Table S5**). Although the model for plasma p-tau181 accounted for 24% of the variability (R^2^=0.24), neither parietal cortex pT181 tau burden, parietal cortex amyloid-β burden, LATE-NC stage, nor Kalaria cerebrovascular disease score significantly contributed when modeled together (**Fig. 3**). In contrast, 51% of the variability in p-tau217 (R^2^=0.51) was accounted for by a similar model. Amyloid-β burden in the parietal cortex (β-coefficient=0.077 [0.026, 0.13], p=0.004) remained the main predictor of p-tau217, but there was not an independent contribution from parietal cortex pT217 tau burden, LATE-NC stage, or Kalaria cerebrovascular disease score.

**Figure 3.**
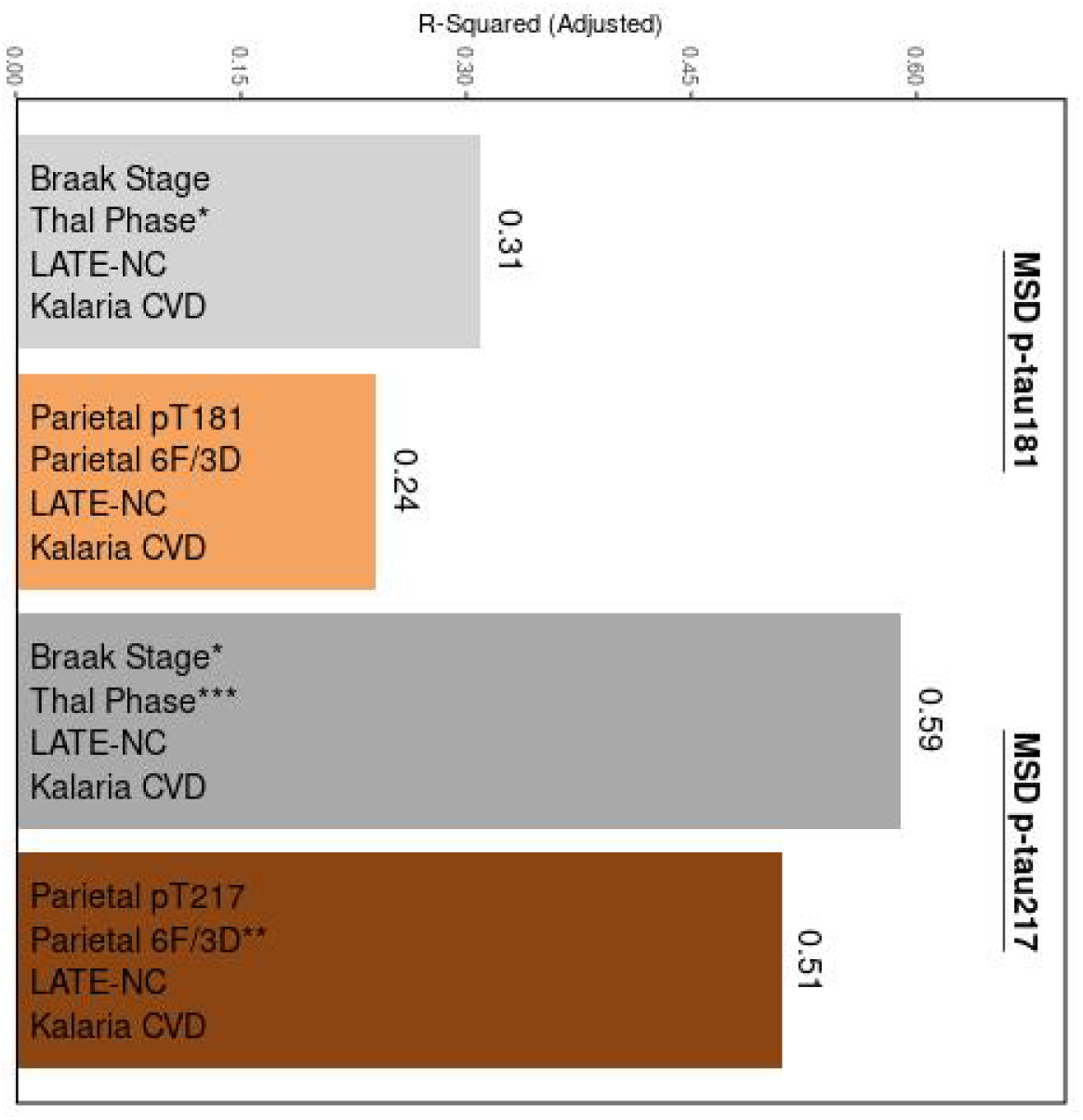
Multivariable linear regression modeling of neuropathologic variables as predictors of plasma p-tau levels. Global scales of tau and amyloid-β (Braak [25] and Thal [26], gray bars) and the strongest regional measure of tau and amyloid-β (parietal cortex, brown bars) were investigated as predictors of variability observed in plasma p-tau181 (left) and p-tau217 levels (right). To account for common co-pathologies, LATE-NC [16] and Kalaria cerebrovascular disease [15] were added to each model. Overall, the global scales performed better than the regional cortical measures with amyloid-β observed as strongest contributor to plasma p-tau variability. Time from plasma draw to death was not used to adjust, as it was not observed to associate with plasma p-tau levels. Case with high creatinine was not included. All variables in the model are shown, corresponding to **Table S5**. Significance denoted as *p<0.05, **p<0.01, ***p<0.001. Acronyms: 6F/3D=amyloid-β antibody. CVD=cerebrovascular disease. LATE-NC=limbic predominant age-related TDP-43 encephalopathy neuropathologic change. MSD=meso scale discovery. p-tau=phosphorylated tau for plasma levels. pT=phosphorylated threonine for immunohistochemical measures of tau.

### Quantitative assessment of neurotransmitter hubs association with plasma p-tau

Based on the strength of the relationship observed between amyloid-β neuropathology and plasma p-tau levels (**Table 2**), along with the diminished relationship with Braak stage (**Table S5**), we next explored the neurotransmitter hubs considered to influence amyloid-β (locus coeruleus [43, 44]) and tau pathology (nucleus basalis of Meynert [45, 46]) through their widespread cortical projections. Nucleus basalis of Meynert neuron count/mm^2^ was not associated with either plasma p-tau181 or plasma p-tau217 (**Table 2, Fig. S7**). In contrast, a lower locus coeruleus neuron count/mm^2^ was strongly associated with higher plasma p-tau181 (R=-0.50, p=0.007) and plasma p-tau217 (R=-0.55, p=0.002).

### Global neuropathologic scales and plasma p-tau evaluated as predictors of cognitive decline

Following assessment of antemortem and postmortem predictors of plasma p-tau variability, we examined the relationship between neuropathology and widely used cognitive measures (CDR [18], MMSE [19]) for generalizability. Based on consistent reports that plasma p-tau levels associated with cognitive decline [1, 3, 8, 9, 47, 48], we sought to investigate whether soluble measures of plasma p-tau independently predicted cognitive decline when accounting for insoluble measures of amyloid-β and tau pathology. To do this we performed linear regression analysis on CDR and MMSE assessed nearest the time of death (**Table S6**) while controlling for time from plasma draw to death. Variability in cognitive scores was similarly predicted for CDR (p-tau181 model: Adj. R^2^=0.25; p-tau217 model: Adj. R^2^=0.27) and MMSE (p-tau181 model: Adj. R^2^=0.30; p-tau217 model: Adj. R^2^=0.32); however, neither was predicted independently by plasma p-tau levels or Thal phase. Braak stage remained the significant predictor of cognitive measures for three of the four models. In the CDR model including plasma p-tau181, for every increase in Braak stage the model predicted 1 point higher on CDR sum of boxes (β-coefficient=1.091 [0.024, 2.158], p=0.045). In the MMSE model including p-tau181, for every increase in Braak stage the model predicted nearly 1 point lower on MMSE (β-coefficient=-0.760 [-1.354, -0.166], p=0.014). The strongest model observed accounted for 32% of variability in MMSE and included plasma p-tau217, which found that for every increase in Braak stage the model predicted nearly 1 point lower on MMSE (β-coefficient=-0.667 [-1.293, -0.040], p=0.038).

## Discussion

In this small series of 35 population-based autopsies from the Mayo Clinic Study of Aging, we report as much as 59% of the variability in plasma p-tau217 can be predicted by a combination of global neuropathologic scales. Tau and amyloid-β neuropathology remained significant predictors of plasma p-tau217, but not severity of LATE-NC stage or cerebrovascular disease scaled score. With as much as 31% of the variability accounted for in plasma p-tau181 levels by global neuropathologic scales, only amyloid-β remained the significant predictor. A common observation across each of the analyses performed revealed plasma p-tau217 to have a stronger association with AD neuropathologic scales than p-tau181.

The neuropathologic findings in this study support the hypothesis that both tau and amyloid-β neuropathology intersect in their influence on plasma p-tau levels (**Fig. 4**). Interestingly, our study and others’ provide supportive evidence that amyloid-β neuropathology has a stronger association with plasma p-tau levels than tau neuropathology [3, 8, 39]. To further extend the detailed work of Wennström and colleagues [49] that focused on the relationship between tau neuropathology and p-tau217, we compared both amyloid-β and tau neuropathology to plasma p-tau181 and p-tau217. Visual examination of this relationship using Thal phase [26] as a global amyloid-β scale demonstrated a biological influence potentially initiating between Thal phase 3 and Thal phase 4, suggesting either neuroanatomic extent to the brainstem or perhaps severity of amyloid-β plaque pathology may be required to cross a threshold. The relationship between regional amyloid-β burden measures from the parietal cortex was weaker than global Thal phase and did not indicate a linear increase in plasma p-tau levels as amyloid-β burden continued to increase in AD cases. Although amyloid-β plaque accumulation does not associate with cognitive decline when accounting for tau pathology [50, 51], supportive evidence suggests neuronal hyperactivation is mediated through an amyloid-β linked defect in synaptic transmission [52, 53]. Increased neuronal activity was previously shown to stimulate the release of tau in vitro, enhance tau pathology in vivo, and lead to production of amyloid-β [54, 55]. We speculate that amyloid-β induced hyperactivation of neurons may impact neuronal dysfunction sufficient to influence release of p-tau into the fluids without enough damage to impact cognitive decline.

**Figure 4.**
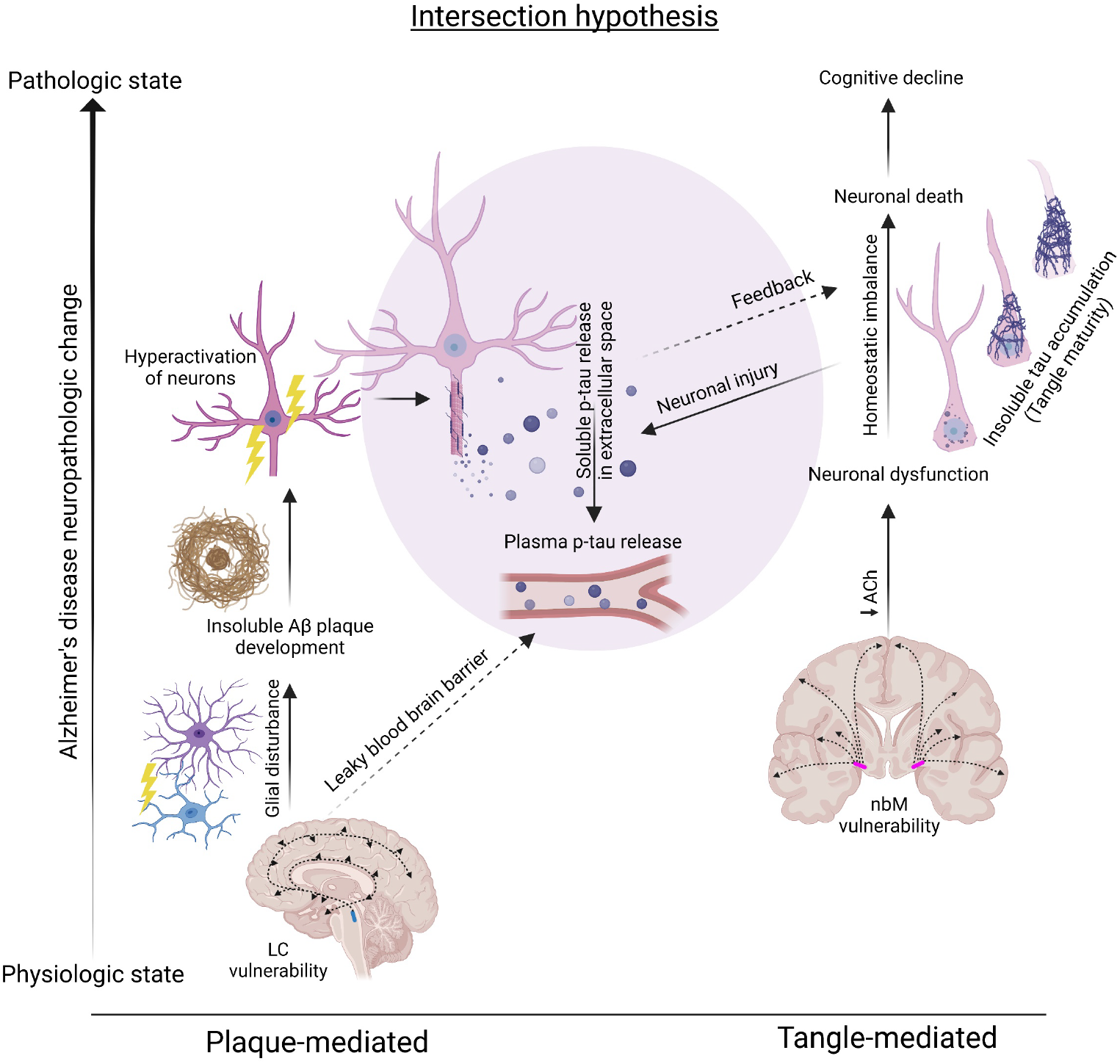
Hypothesized intersection of amyloid-β plaque and neurofibrillary tangle pathology in cortex and their influence on soluble p-tau release into plasma. We propose parallel processes occur for amyloid-β plaque (**Left**) and neurofibrillary tangle (**Right**) pathologies with an intersection (**Middle**) in the AD brain that impacts phospho-tau release into plasma. The hypothesized intersection is influenced by data from the cortex and may not apply to limbic regions, as the current study and others [49] observed a weaker relationship between limbic neuropathology and plasma p-tau levels. **Plaque-mediated route:** The locus coeruleus is highly vulnerable in the AD brain and is considered the earliest site of phospho-tau accumulation [68]. This noradrenergic hub nucleus (mid-sagittal brain, blue spot) sends projections throughout the brain [43] and is thought to influence amyloid-β plaque deposition through a mechanistic effect on glial disturbance [44]. Based upon available evidence that amyloid-β deposition induces hyperactivation of neurons [52, 53], we speculate that the plaque-mediated impact on soluble p-tau release is a the result of increased neuronal activity that stimulates and enhances release of p-tau into the extracellular space [54, 55]. The association observed in the current study between lower locus coeruleus neuron counts and higher plasma p-tau levels is thought to be hypothesized to be the result of noradrenergic deficiencies leading to increased leakage of blood-brain barrier [60] enhancing release of p-tau into plasma. **Tangle-mediated route:** The nucleus basalis of Meynert acts as a cholinergic hub (coronal brain, pink spot) that supplies acetylcholine to the cortex and is thought to influence cortical tangle accumulation through cholinergic deficiencies leading to neuronal dysfunction [45, 46, 61]. We did not observe an association between nucleus basalis of Meynert neuron count and plasma p-tau levels, which may suggest the resultant neuronal dysfunction from decreased acetylcholine does not affect p-tau release. The current study and others [4, 49] observed a strong relationship between severity of neurofibrillary tangle accumulation and plasma p-tau, which we speculate may result from neuronal injury [69] as tangles mature through their lifespan [70, 71]. Amyloid-β plaque accumulation does not associate with cognitive decline when accounting for tau pathology [50, 51], however we and others previously showed plasma p-tau to associate with cognitive decline [1, 3, 8, 9, 47, 48]. To evaluate both neuropathologic and soluble plasma p-tau contribution to cognitive decline, the current study utilized regression modeling. The results suggest that underlying tangle accumulation in the brain (Braak stage), but neither amyloid-β deposition (Thal phase) nor soluble plasma p-tau levels predict cognitive decline when modeled simultaneously. Acronyms: Aβ=amyloid-β. AD= Alzheimer’s disease. LC=locus coeruleus. nbM=nucleus basalis of Meynert. p-tau=phosphorylated tau. T=threonine. Created with BioRender.com

Given the strength of the relationship between amyloid-β and plasma p-tau levels, we explored the novel hypothesis that neuronal loss in the locus coeruleus would associate with higher plasma p-tau levels. The locus coeruleus is a noradrenergic hub nucleus that sends projections throughout the brain and is thought to influence amyloid-β plaque deposition through a mechanistic effect on glial disturbance [43, 44]. Even in our small unselected series we observed a strong relationship between neuronal loss in the locus coeruleus and higher p-tau181 and p-tau217 levels. The locus coeruleus is an area observed to develop neurofibrillary tangle pathology prior to entorhinal cortex [37, 56] that is currently being evaluated as an early AD biomarker [57, 58]. Furthermore, evidence from animal studies suggests damage to the locus coeruleus may impact cerebrovascular clearance mechanisms [59, 60]. We propose that the resultant increase in microvasculature permeabilty following locus coeruleus damage and amyloid-β-induced hyperexcitabilty of neurons may be the mechanism underlying the relationship between amyloid-β neuropathology and soluble p-tau release into plasma (**Fig. 4**).

Although we and others previously demonstrated a relationship between nucleus basalis of Meynert neuronal loss and cortical vulnerability to tangle pathology in AD [45, 46, 61, 62], we did not observe a relationship with p-tau plasma levels. We speculate that neuronal dysfunction related to impaired cholinergic projections may be insufficient to influence release of soluble p-tau into plasma. We did observe an association between Braak stage, a widely used global scale of tau pathology in the brain [25], and plasma p-tau levels. This confirms several reports demonstrating this robust association [9, 11, 39, 63]. A discernible threshold in p-tau levels was observed between Braak III and Braak IV, which corresponds with accumulation of cortical tau pathology by Braak IV [25, 37]. To further examine a more direct relationship with cortical severity, we immunostained tissue sections with pT181 and pT217 antibodies that recognize the same phosphorylation sites studied in plasma. Increasing immunohistochemical burden for both tau antibodies in AD cases appeared to have a ceiling effect without the observation of a linear increase in plasma p-tau levels. This supports data from a longitudinal study that stratified cases by Braak stage and observed a plasma p-tau181 ceiling effect [64]. Regional cortical tau burden had a stronger relationship with plasma p-tau levels than global Braak stage, which may reflect greater power to detect differences using quantitative methods over semi-quantitative global scales.

The association of hippocampal tau burden with both plasma p-tau181 and p-tau217 levels was modest in comparison to the strength of the relationship observed with either cortical tau or Braak stage. This supports prior work demonstrating higher correlation with cortical tau measures compared to medial temporal lobe structures using pT217 antibodies [49], and further confirms this relationship with pT181. As the hippocampus is a small region, p-tau levels may better reflect global involvement overrepresented by cortical tau load. An additional consideration for the weakened relationship in the hippocampus is that age-related tau accumulation in limbic structures that occurs independent of amyloid-β (i.e., primary age-related tauopathy [29]) may be insufficient to affect a discernible release of soluble p-tau into plasma [39]. We previously demonstrated that pT181 and pT217 antibodies recognize earlier aspects of tangle maturity [35], including pretangles and mature tangles. Thus, early tangle accumulation in the cortex may be more readily detected by the selected antibodies, enabling the observed relationship to be detected. In comparison, the hippocampus may have long-standing advanced tangle accumulation not as readily captured by the pT181 and pT217 antibodies.

We consistently observed a stronger relationship between neuropathologic measures and plasma p-tau217, compared to p-tau181. Moreover, the ratio between hippocampus to parietal cortex pT181 burden was muted in comparison to that observed in pT217. One possible explanation may be differences in antibody recognition of physiologic versus pathologic p-tau. Although uncommon, there were cases excluded from regional pT181 analyses as the immunohistochemical burden reflected axonal staining rather than tau pathology. A high-resolution quantitative proteomics study of tau demonstrated that phosphorylation at threonine-181 was observed in 70% of clinical controls with a Braak stage <IV and 92% of AD cases [65]. In comparison, phosphorylation at threonine-217 was observed in 14% of controls and 86% of AD cases [65]. These observations of physiologic differences between p-tau181 and p-tau217 should not detract from the prognostic data on p-tau181 [4, 6, 7, 9-11, 63], but may provide a deeper understanding of why p-tau217 associates more specifically with neuropathologic accumulation of tau pathology in the human brain.

Our study has several limitations that are important to consider, namely the small sample size. Although we investigated population-based study participants, all 35 participants who came to autopsy with plasma p-tau levels within 3 years were non-Hispanic white. With our finding that kidney health may impact plasma p-tau levels, further investigation in ethnoracially diverse cohorts remains critical as the prevalence of chronic kidney disease varies [66]. While our series had a range of neuropathologic diagnoses, small sample sizes precluded group comparisons. To offset this limitation, graphs are presented with visual representation of disease groups for interpretation of AD neuropathologic change (i.e., AD, pathological aging) in comparison with primary tauopathies (i.e., primary age-related tauopathy, argyrophilic grain disease, and progressive supranuclear palsy). To expand our understanding between neuropathologic changes and plasma p-tau levels, we utilized digital pathology methods. As a result, the 6F/3D amyloid-β antibody was used for evaluation as the 6E10 antibody labels intracellular APP and was expected to interfere with burden analysis. However, it should be noted that in some cases the 4G8 antibody may be more sensitive to diffuse plaque pathology [26, 67], which could have an effect on performing Thal phase with 6F/3D. Our quantitative digital pathology studies were only performed in the CA1-subiculum of the hippocampus and parietal cortex, which may limit interpretation with other brain regions. Most of our cases were older than 75 years at death, therefore limiting our interpretation in younger autopsy cohorts. We did not observe a relationship between age at plasma draw and plasma p-tau levels, but more work is needed in younger cases to see if this remains a consistent finding.

## Conclusions

Our findings provide strong evidence that soluble plasma p-tau levels reflect insoluble accumulation of amyloid-β and tau pathology. As cut-points for diagnostic utility continue to undergo determination, it is important to note that plasma p-tau negativity does not exclude for the possibility of underlying tau and amyloid-β pathology. Instead, plasma p-tau positivity will reflect a threshold crossed where p-tau levels correspond to a functional measure ascribed to significant disease-relevant changes. We propose parallel processes occur for amyloid-β deposits and tangle development with an intersection between these neuropathologies in AD that impacts soluble p-tau release, but not necessarily cortical tangle accumulation. The association observed between locus coeruleus and plasma p-tau levels suggests noradrenergic deficiencies may play a role in release of tau that we hypothesize to be mediated through amyloid-β-induced hyperactivation of neurons and increased microvasculature leakage.

## Data Availability

All data produced in the present study are available upon reasonable request to the authors.

## Declarations

### Ethics approval and consent to participate

The Mayo Clinic and Olmsted Medical Center Institutional Review Boards approved this study. All participants provided written informed consent.

### Consent for publication

Not applicable

### Availability of data and materials

All requests for raw and analyzed data and related materials will be reviewed by Mayo Clinic’s Legal Department and Mayo Clinic Ventures to verify whether each request is subject to any intellectual property or confidentiality obligations. Requests for patient-related data not included in the paper will not be considered. Any data and materials that can be shared will be released via a Data Use/Share Agreement or Material Transfer Agreement.

## Author Contributors

MEM and MMM designed the study and performed primary interpretation of the data. MEM wrote the manuscript, with MMM, JLD, and CRJ Jr. contributing substantial edits. RCP, CRJ Jr., DSK, PV, JGR were responsible for acquisition of clinical data. MEM, ATN, RRR, and DWD were responsible for acquisition of neuropathology data. MEM, CMM, DMR, JFT, TNHS were responsible for neurohistology organization and digital pathology analyses. MEM, BDCB, JLD, MMM were responsible for interpretation of plasma p-tau data. NK performed Kalaria cerebrovascular disease scoring. NK and BJM evaluated TDP-43 pathology. ATN, RRR, and DWD were responsible for neuropathologic examination. JAS performed statistical analyses. Funding was obtained by MEM, RCP, PV, JGR, MMM. All authors contributed and critically reviewed the final version of the manuscript. All authors read and approved the final manuscript.

## Acknowledgements

We thank Monica Castanedes-Casey, Nathan Perez, and Virginia Phillips for histologic services. Programmatic support by Sabrina Rothberg and Kelsey Caetano-Anolles continues to be invaluable, and we are appreciative of their dedication. We are grateful to the patients and families for their generous brain donations to help further our knowledge of Alzheimer’s disease.

## Abbreviations

AD: Alzheimer’s disease
AGD: argyrophilic grain disease
ALT: alanine transaminase
AST: aspartate aminotransferase
AUC: area under the curves
CDR: Clinical Dementia Rating
CERAD: Consortium to Establish a Registry for Alzheimer’s Disease
IQR: interquartile range
LATE-NC: limbic-predominant age-related TDP-43 encephalopathy neuropathologic change
MCSA: Mayo Clinic Study of Aging
MMSE: Mini-Mental State Examination
MSD: meso scale discovery
PART: primary age-related tauopathy
PET: positron emission tomography
p-tau: phosphorylated-tau
p-tau181: phosphorylated-tau at threonine181
p-tau217: phosphorylated-tau at threonine217
REP: Rochester Epidemiology Project
ROC: receiver operating characteristic
SUVr: standardized uptake value ratios
TDP-43: TAR DNA binding protein 43

## Supplementary Appendix

### Supplemental Methods

#### Plasma p-tau assays

Phosphorylated tau at threonine181 (p-tau181) and threonine217 (p-tau217) levels were measured in duplicate on a streptavidin small spot plate using the meso scale discovery (MSD) platform by electrochemiluminescence using proprietary assays developed by Lilly Research Laboratories, as previously described [1]. Briefly, samples were diluted 1:2 and 50 uL of diluted sample was used for each replicate. P-tau181 used biotinylated-AT270 (mouse IgG1) as the capture and p-tau217 used biotinylated-IBA493 (mouse IgG1) as the capture. In this study, both assays used SULFO-4G10-E2 (anti-tau monoclonal antibody developed by Lilly Research Laboratories) as the detector. Each assay was calibrated using a unique synthetic p-tau peptide coupled with a polyethylene glycol linker to a second tau peptide matching amino acid 111-130 according to the Tau441 sequence numbering.

#### Digital pathology procedures

Hematoxylin and eosin (H&E), as well as immunohistochemically stained slides, were digitally scanned at 20x using the Aperio AT2 scanner and annotated using ImageScope (Leica Biosystems, version 12.4.3.5008). The CA1 and subiculum subsectors of the hippocampus were traced on serial sections as previously described [2]. Briefly, the superior border was defined as the boundary between the lacunosum and radiatum layers. The inferior border was defined as the boundary between the pyramidal layer and alveus. To operationalize using neuroanatomic landmarks, the CA1 and subiculum were collectively traced from the ventricle following the alveus medially to the rise of the presubiculum. The inferior parietal cortex was traced using the pial surface and gray-white junction as borders along the extent of the gray matter. Given the lack of discrete boundaries in the locus coeruleus, a 1.8×1.8 mm^2^ box was placed centering the locus coeruleus neurons based upon maximization of locus coeruleus neurons encompassed in 10 healthy control brains. The level (i.e. rostral-to-caudal extent) of the locus coeruleus was assessed using appearance of the superior cerebellar peduncle, velum, cerebral aqueduct, and fourth ventricle (**Fig. S2**) [3]. The middle locus coeruleus level was selected for analysis based on prior studies demonstrating topographic vulnerability of rostral and middle level to AD [3, 4] and wider availability in the study cohort (n=28/35 [80%]). The nucleus basalis of Meynert was annotated as previously described [5]. Briefly, the lateral edge of the nucleus basalis of Meynert was defined by where the globus pallidus and putamen meet perpendicular to the ventral surface of the brain. The medial edge was defined by a perpendicular line from the lateral end of the optic nerve up to the globus pallidus, or the most medial part of the internal capsule if the optic nerve was not present. The anterior commissure was dorsal to the annotated region. The nucleus basalis of Meynert level (i.e., anterior-to-posterior extent) was assigned using the location and appearance of the anterior commissure, globus pallidus, fornix, mammillary body (**Fig. S3**). The anterior nucleus basalis of Meynert level was selected for analysis because this level contains the most widespread and noticeable portion of the nucleus basalis of Meynert [5-7], along with wider availability in the study cohort (n=24/35 [69%]).

GENIE, a pattern recognition software (Leica Biosystems, v12.4.3.7001) was used to better automate locus coeruleus and nucleus basalis of Meynert neuron counts (**Fig. S4**). Annotated examples of each classifier were used to train until a mean training accuracy >85% was reached with minimal errors identified visually. For the locus coeruleus, four classifiers were created: locus coeruleus neurons, blood vessels, neuropil, and white matter. After 1,000 iterations of training, a mean training accuracy of 98.28% was reached. The classifiers were then incorporated into a nuclear macro so that regions classified as “LC neurons” and “neuropil” would be analyzed. For the nucleus basalis of Meynert, five classifiers were created: nucleus basalis of Meynert neurons, blood vessels, neuropil, corpora amylacea, and small vessels. After 2,000 iterations of training, a mean training accuracy of 88.19% was reached. The classifiers were then incorporated into a nuclear macro so that regions classified as “nbM neurons” and “neuropil” would be analyzed. eSlideManager (Leica Biosystems) was used to analyze digital images of the hippocampus and parietal cortex. Slides were batch analyzed using customized color deconvolution and positive pixel count macros designed for each antibody to recognize the 3, 3’-diaminobenzidine staining on the tissue and to exclude background to obtain burden (**Tables S2-S3**). Scans of the locus coeruleus and nucleus basalis of Meynert batch analyzed using customized nuclear macros designed for each region, including their GENIE classifiers (**Table S4**).

#### Tau PET procedures

To contextualize the strength of the relationship between regional tau measures and plasma p-tau levels, antemortem tau positron emission tomography (PET) uptake was quantified in inferior parietal cortex for 10 cases with available data (**Table S7**). Tau PET measures from hippocampus are often difficult to ascertain and were thus not investigated. [^18^F]Flortaucipir tau PET images were acquired using a PET/CT scanner operating in 3-dimensional mode, as previously described in detail [8]. Inferior parietal cortex region of interest (ROI) was defined by an in-house version of the automated anatomic labelling atlas as previously described. The tau PET ROI median inferior parietal cortex values were normalized to cerebellar crus (bilateral crus, 1-2) to calculate regional standardized uptake value ratios (SUVr) [8].

### Supplemental Results

#### Clinical diagnosis across autopsy cohort

Autopsied participants included cognitively unimpaired (n=25 [71%]), mild cognitive impairment (n=4 [11%]), AD dementia (n=3 [9%]), and non-AD neurodegenerative disorders (n=3 [9%]) (**Table S7**).

#### Antemortem contributors to plasma p-tau levels

Because previous studies suggested that comorbidities, including both kidney and liver disease, may affect p-tau biomarker levels,[9, 10] we first examined Spearman correlations of creatinine, aspartate aminotransferase (AST), and alanine aminotransferase (ALT) with plasma p-tau levels. Graphs of creatinine and plasma p-tau levels are shown in **Figure S5-A, F**. Serum creatinine levels correlated with plasma p-tau181 (R=0.34, p=0.049) and approached significance with plasma p-tau217 (R=0.31, p=0.074). One individual was an outlier with high serum creatinine (3.7 mg/dL). This person also had the highest plasma p-tau181 (10 pg/mL) and plasma p-tau217 (1.3 pg/mL) levels of the cohort. Neuropathologic investigation of the brain of this man in his 90s revealed argyrophilic grains disease and pathological aging (Braak III, Thal 3), which was deemed insufficient to account for the high plasma p-tau levels. Exclusion of the outlier diminished the relationship between creatinine and p-tau181 (R=0.27, p=0.116), as well as plasma p-tau217 (R=0.24, p=0.169). This individual remained part of the initial analyses evaluating antemortem variability of creatinine, AST, ALT, age at plasma p-tau draw, and time from blood draw to death, but was removed from subsequent analyses investigating cognition and neuropathology. We next examined liver health with a focus on serum AST (**Figure S5-B, G**) and ALT liver enzyme levels (**Figure S5-C, H**). Serum AST levels did not correlate with plasma p-tau181 (R=-0.21, p=0.257) or with plasma p-tau217 (R=-0.12, p=0.526). Similarly, we did not observe a correlation between serum ALT levels and plasma p-tau181 (R=0.14, p=0.553) or with plasma p-tau217 (R=0.14, p=0.564). The male participant in his 90s with high serum creatinine levels (3.7 mg/dL) had the shortest time from plasma draw to death (0.3 years). Exclusion of this participant did not improve the relationship between either age at plasma draw or time from plasma draw to death with plasma p-tau levels.

#### Tau PET signal in parietal cortex associates with plasma p-tau levels

A subset of 10 individuals with antemortem [^18^F]flortaucipir PET were examined for their relationship between radioligand uptake in parietal cortex and plasma p-tau levels to provide in vivo comparison (**Table S7, Fig. S8**). The magnitude of the correlation between parietal cortex tau PET uptake and plasma p-tau181 was high, but did not reach statistical significance (R=0.54, p=0.113). We observed an even stronger relationship between parietal cortex tau PET uptake and plasma p-tau217 (R=0.71, p=0.028).

#### Alzheimer’s disease neuropathologic change predicted better by cognitive measures than plasma p-tau

Although we hypothesized that accumulation of neuropathology influences plasma p-tau levels, we evaluated receiver operating characteristics (ROC) curves with corresponding area under the curves (AUC) to assess the utility of plasma p-tau and cognitive scores to independently prognosticate neuropathologic classification of intermediate-to-high from none-to-low AD neuropathologic change (**Fig. S9**). Given the sample size and use of logistic regression, plasma p-tau levels and cognitive scores were modeled individually without covariates. P-tau217 had the highest predictive value for AD neuropathologic change (AUC=0.89 [CI=0.75, 1.0]), followed by CDR sum of boxes (AUC=0.81 [0.66, 0.95]), p-tau181 (AUC=0.80 [CI=0.62, 0.95]), and MMSE (AUC=0.76 [CI=0.56, 0.93]).

### Supplemental Tables

**Table S1.**
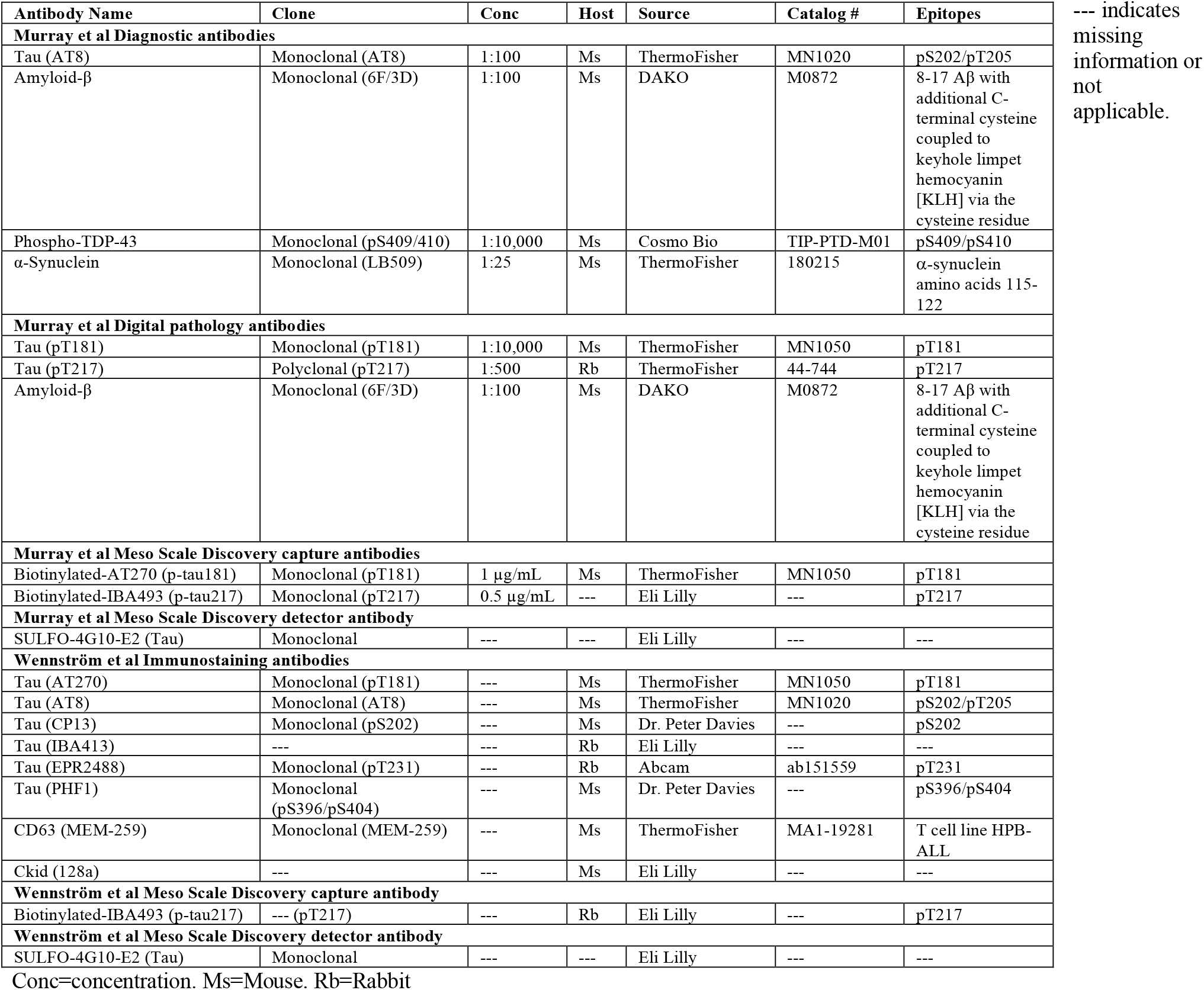
Antibody table.

**Table S2.** Digital pathology color deconvolution count macros for pT181 and pT217.

|  |  |
| --- | --- |
| **Version** | v9 |
| **View Width** | 1000 |
| **View Height** | 1000 |
| **Overlap Size** | 0 |
| **Image Zoom** | 1 |
| **Markup Compression Type** | Same as processed image |
| **Compression Quality** | 30 |
| **Classifier Neighborhood** | 0 |
| **Classifier** | None |
| **Class List** |  |
| **Positive Color Channel** | 3 |
| **Mark-up Image Type** | Intensity Ranges |
| **Weak Positive Threshold** | 235 |
| **Medium Positive Threshold** | 180 |
| **Strong Positive Threshold** | 180 |
| **Black Threshold** | 0 |
| **Color (1) - Red Component** | 0.65 |
| **Color (1) - Green Component** | 0.704 |
| **Color (1) - Blue Component** | 0.286 |
| **Color (2) - Red Component** | 0.072 |
| **Color (2) - Green Component** | 0.99 |
| **Color (2) - Blue Component** | 0.105 |
| **Color (3) - Red Component** | 0.341 |
| **Color (3) - Green Component** | 0.575 |
| **Color (3) - Blue Component** | 0.743 |
| **Clear Area Intensity** | 240 |
| **Display Plots** | No |

**Table S3.** Positive pixel count macro specifications for 6F/3D.

|  |  |
| --- | --- |
| **Version** | **v9** |
| **View Width** | 1000 |
| **View Height** | 1000 |
| **Overlap Size** | 0 |
| **Image Zoom** | 1 |
| **Markup Compression Type** | Same as processed image |
| **Compression Quality** | 30 |
| **Classifier Neighborhood** | 0 |
| **Classifier** | None |
| **Class List** |  |
| **Hue Value** | 0.1 |
| **Hue Width** | 0.1 |
| **Color Saturation Threshold** | 0.08 |
| **Iwp (High)** | 235 |
| **Iwp(Low) = Ip(High)** | 235 |
| **Ip(Low) = Isp(High)** | 235 |
| **Isp(Low)** | 0 |
| **Inp(High)** | -1 |

**Table S4.**
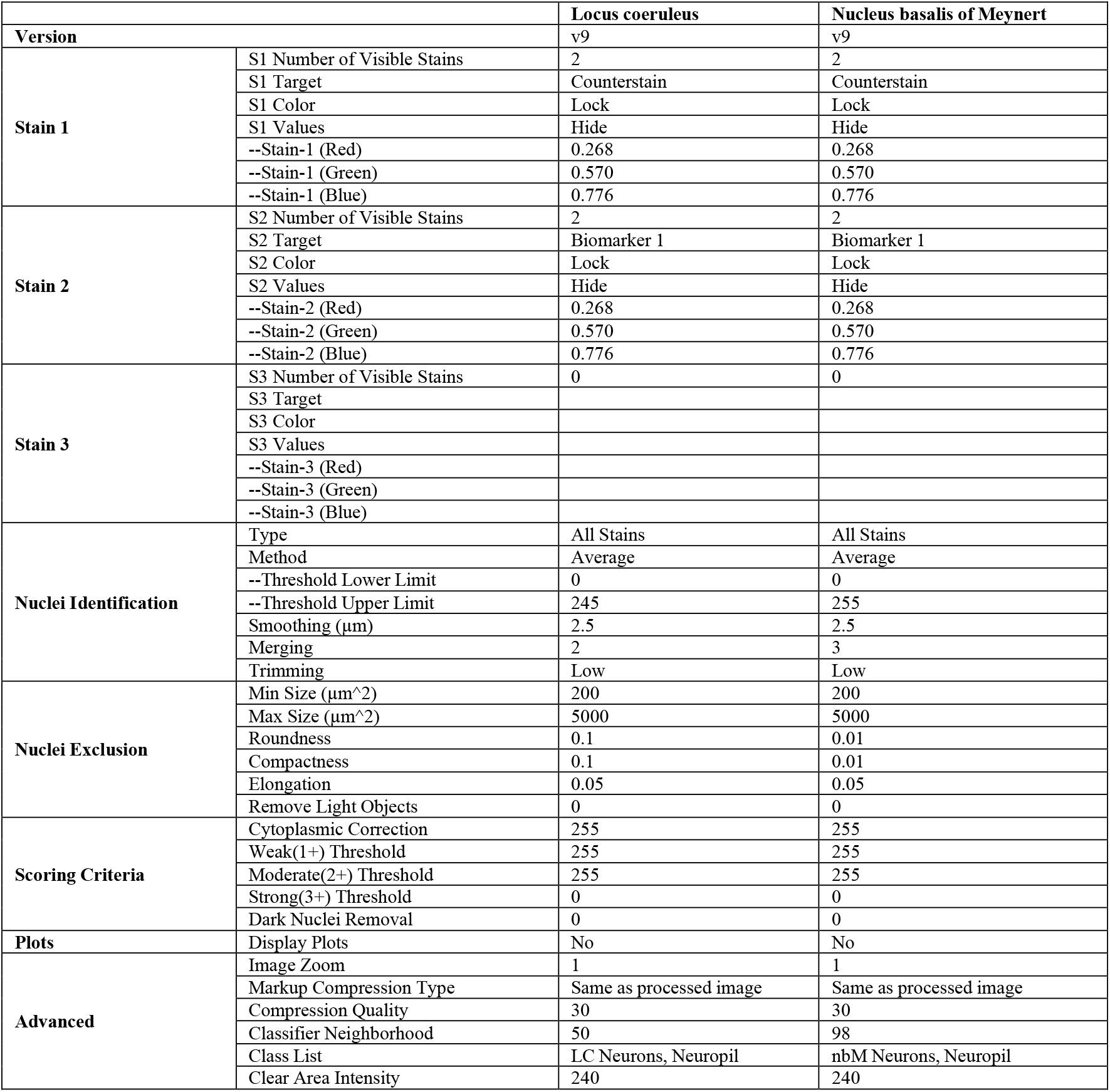
Nuclear macros for digital pathology.

**Table S5:** Multivariable linear regression models investigating contribution of co-existing pathology to the associations between global scales and regional neuropathologic measures to plasma p-tau levels.

| Tau & Amyloid-β | Outcome | Adj R<sup>2</sup> | Predictor | β-coefficient | LowerCL | UpperCL | p-value |
| --- | --- | --- | --- | --- | --- | --- | --- |
| **Global** | MSD p-tau181 | 0.31 | Braak stage | 0.2681 | -0.0726 | 0.6089 | 0.1184 |
|  |  |  | Thal phase | 0.3269 | 0.0528 | 0.6010 | 0.0211 |
|  |  |  | LATE-NC stage | 0.0051 | -0.5008 | 0.5109 | 0.9838 |
|  |  |  | Kalaria CVD score | 0.0111 | -0.1622 | 0.1844 | 0.8966 |
|  | MSD p-tau217 | 0.59 | Braak stage | 0.0601 | 0.0120 | 0.1081 | 0.0160 |
|  |  |  | Thal phase | 0.0803 | 0.0417 | 0.1190 | 0.0002 |
|  |  |  | LATE-NC stage | 0.0132 | -0.0581 | 0.0845 | 0.7085 |
|  |  |  | Kalaria CVD score | -0.0039 | -0.0284 | 0.0205 | 0.7442 |
| **Regional** | MSD p-tau181 | 0.24 | Parietal pT181 | 0.6620 | -0.5770 | 1.9010 | 0.2804 |
|  |  |  | Parietal 6F3D | 0.2678 | -0.0560 | 0.5916 | 0.1005 |
|  |  |  | LATE-NC stage | 0.1171 | -0.4697 | 0.7039 | 0.6835 |
|  |  |  | Kalaria CVD score | -0.0372 | -0.2637 | 0.1894 | 0.7374 |
|  | MSD p-tau217 | 0.51 | Parietal pT217 | 0.0217 | -0.0222 | 0.0656 | 0.3209 |
|  |  |  | Parietal 6F3D | 0.0766 | 0.0262 | 0.1270 | 0.0042 |
|  |  |  | LATE-NC stage | 0.0250 | -0.0526 | 0.1027 | 0.5151 |
|  |  |  | Kalaria CVD score | 0.0072 | -0.0183 | 0.0327 | 0.5683 |

Analyses were performed on n=34 individuals for p-tau217. Analyses were performed on n=28 individuals for p-tau181 as a result of immunohistochemical exclusion in 5 cases with heavy pT181 staining appearing in axons disproportionate to tau pathology. Case with high creatinine was not included. All variables in model are shown. Time from plasma draw to death was not used to adjust, as it was not observed to associate with plasma p-tau levels. Variance inflation factor for all terms were below 3, which fell below the general cutoff of 5 used to indicate multicollinearity. Acronyms: 6F/3D=amyloid-β antibody. Adj=adjusted, CL=95% confidence limit. Acronyms: CVD=cerebrovascular disease. LATE-NC=limbic predominant age-related TDP-43 encephalopathy neuropathologic change. MSD=meso scale discovery. pT=phosphorylated threonine for immunohistochemical measures of tau. p-tau=phosphorylated tau for plasma levels.

**Table S6:** Multivariable linear regression models investigating neuropathologic and plasma p-tau predictors of cognitive scores.

| Cognitive test | Outcome | Adj R<sup>2</sup> | Predictor | β-coefficient | LowerCL | UpperCL | p-value |
| --- | --- | --- | --- | --- | --- | --- | --- |
| CDR Sum of Boxes | CDR Sum of Boxes | 0.25 | MSD p-tau181 | 0.1079 | -1.0906 | 1.3065 | 0.8551 |
|  |  |  | Braak stage | 1.0912 | 0.0243 | 2.1581 | 0.0453 |
|  |  |  | Thal phase | 0.4581 | -0.4722 | 1.3884 | 0.3222 |
|  |  |  | Plasma to death | 1.3458 | -0.6277 | 3.3193 | 0.1737 |
|  | CDR Sum of Boxes | 0.27 | MSD p-tau217 | 3.9035 | -4.4259 | 12.233 | 0.3457 |
|  |  |  | Braak stage | 0.8842 | -0.2347 | 2.0031 | 0.1169 |
|  |  |  | Thal phase | 0.1806 | -0.8857 | 1.2469 | 0.7315 |
|  |  |  | Plasma to death | 1.3052 | -0.6370 | 3.2474 | 0.1798 |
| MMSE | MMSE | 0.30 | MSD p-tau181 | -0.0877 | -0.7665 | 0.5910 | 0.7926 |
|  |  |  | Braak stage | -0.7601 | -1.3537 | -0.1665 | 0.0141 |
|  |  |  | Thal phase | -0.2814 | -0.8001 | 0.2373 | 0.2750 |
|  |  |  | Plasma to death | -0.0450 | -1.1919 | 1.1018 | 0.9363 |
|  | MMSE | 0.32 | MSD p-tau217 | -2.1001 | -7.2021 | 3.0019 | 0.4052 |
|  |  |  | Braak stage | -0.6666 | -1.2934 | -0.0397 | 0.0380 |
|  |  |  | Thal phase | -0.1638 | -0.7536 | 0.4259 | 0.5729 |
|  |  |  | Plasma to death | -0.0800 | -1.2102 | 1.0501 | 0.8854 |

Analyses were performed on n=34 individuals for CDR and n=31 for MMSE. Cognitive test date nearest to death was used to evaluate neuropathologic and plasma p-tau predictors of variability in CDR and MMSE. Although time from plasma draw date to death (measured in years) was not observed to associate with plasma p-tau levels, this time factor was included in the model to adjust for any differences in time lag. Case with high creatinine was not included. All variables in model are shown. Variance inflation factor for all terms were below 3, which fell below the general cutoff of 5 used to indicate multicollinearity. Acronyms: Adj=adjusted. CDR Sum of Boxes=clinical dementia rating sum of boxes. CL=95% confidence limit. Acronyms: MSD=meso scale discovery. MMSE=mini mental state examination. P-tau=phosphorylated tau for plasma levels.

**Table S7.** Clinical diagnosis and availability of tau PET observed within each neuropathologic grouping.

|  | Neuropathologic grouping |  |  |  |  |
| --- | --- | --- | --- | --- | --- |
|  | PSP (n=2) | AD (n=9) | AGD (n=2*) | PART (n=14) | PA (n=7) |
| **Clinical diagnosis** |  |  |  |  |  |
| Cognitively unimpaired, % | 1/2 (50%) | 2/9 (22%) | 1/2 (50%) | 14/14 (100%) | 6/7 (86%) |
| Mild cognitive impairment | 0/2 (0%) | 2/9 (22%) | 1/2 (50%) | 0/14 (0%) | 1/7 (14%) |
| AD dementia | 0/2 (0%) | 3/9 (33%) | 0/2 (0%) | 0/14 (0%) | 0/7 (0%) |
| Non-AD neurodegenerative disorder |  |  |  |  |  |
| Dementia hard to classify | 0/2 (0%) | 2/9 (22%) | 0/2 (0%) | 0/14 (0%) | 0/7 (0%) |
| Progressive supranuclear palsy | 1/2 (50%) | 0/9 (0%) | 0/2 (0%) | 0/14 (0%) | 0/7 (0%) |
| **Tau PET availability** | 0/2 (0%) | 4/9 (44%) | 0/2 (0%) | 5/14 (36%) | 1/7 (14%) |

\*AGD case with high creatinine was not included in table, as the cognitively unimpaired individual was excluded from clinicopathologic analyses after plasma creatinine levels were found to be an outlier influencing plasma p-tau levels. Note: The clinical diagnoses for AD neuropathologic grouping does not sum to 100% due to rounding rules. Acronyms: AD=Alzheimer's disease. AGD=Argyrophilic grains disease. PA=pathological aging. PART=primary age-related tauopathy. PET=positron emission tomography. PSP=progressive supranuclear palsy.

### Supplemental Figures

**Figure S1.**
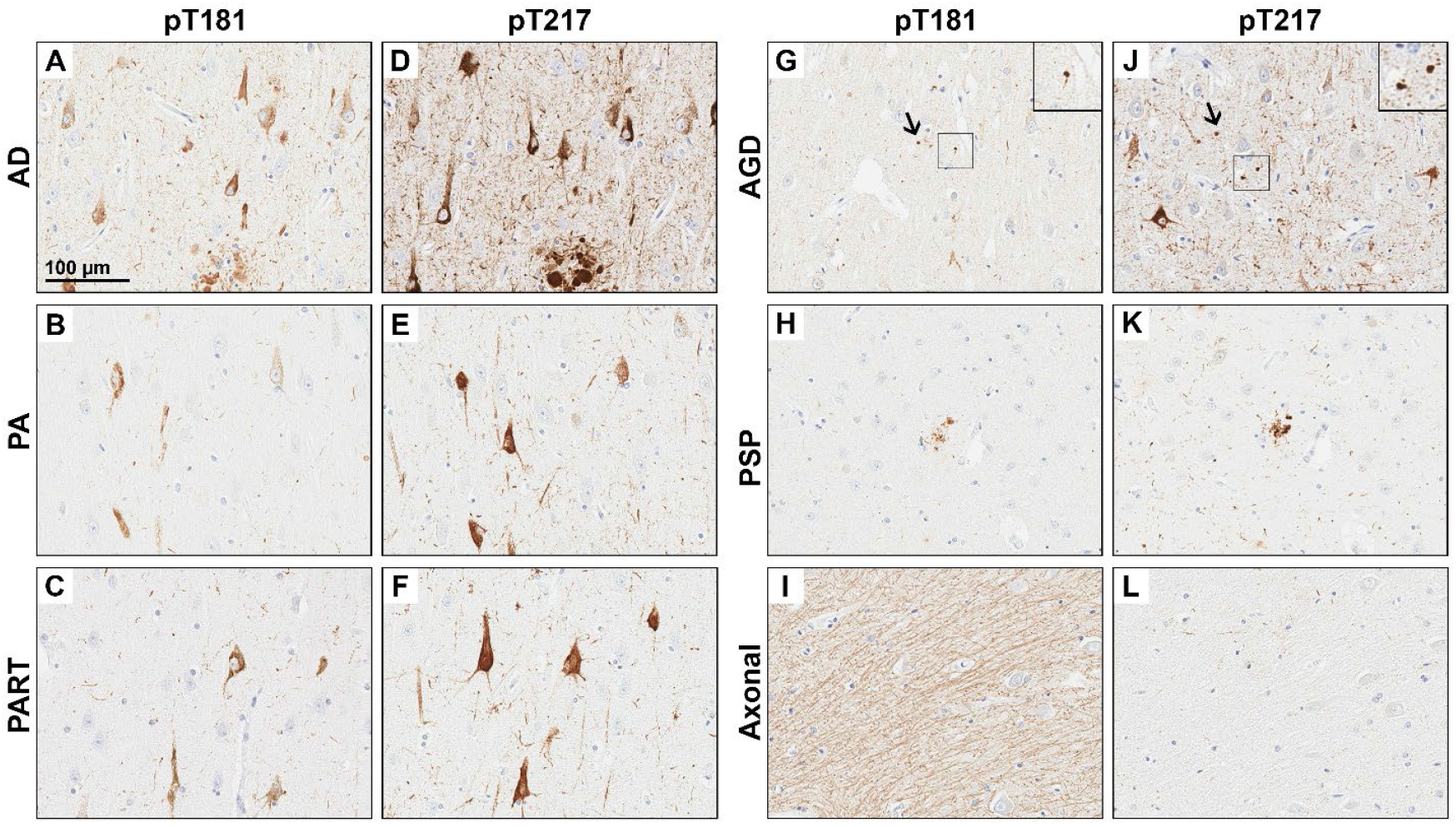
Immunohistochemical staining from hippocampus across disease groups. We commonly observed pT181 **(A-C**,**D-F)** and pT217 **(G-I**,**J-L)** immunostaining in tangle-bearing neurons and neuropil threads in the hippocampus of Alzheimer’s disease (Braak VI, Thal 5) **(A, G)** pathological aging (male in his 80s, Braak II, Thal 3) **(B, H)** primary age related tauopathy (Braak III, Thal 1) **(C, I)**, argyrophilic grain’s disease (Braak II, Thal 0) (**D, J**; grains inset), and progressive supranuclear palsy (Braak III, Thal 3) **(E, K)**. Nonpathologic tau immunostaining of axons (Braak III, Thal 0) was observed in some case on pT181 **(F)**, but not in pT217 **(L)**. Scale bar represents 100 µm. Acronyms: AD=Alzheimer’s disease. AGD=argyrophilic grain disease. PA=pathological aging. PART=primary age related tauopathy. PSP=progressive supranuclear palsy. P-tau=phosphorylated tau for plasma levels.

**Figure S2.**
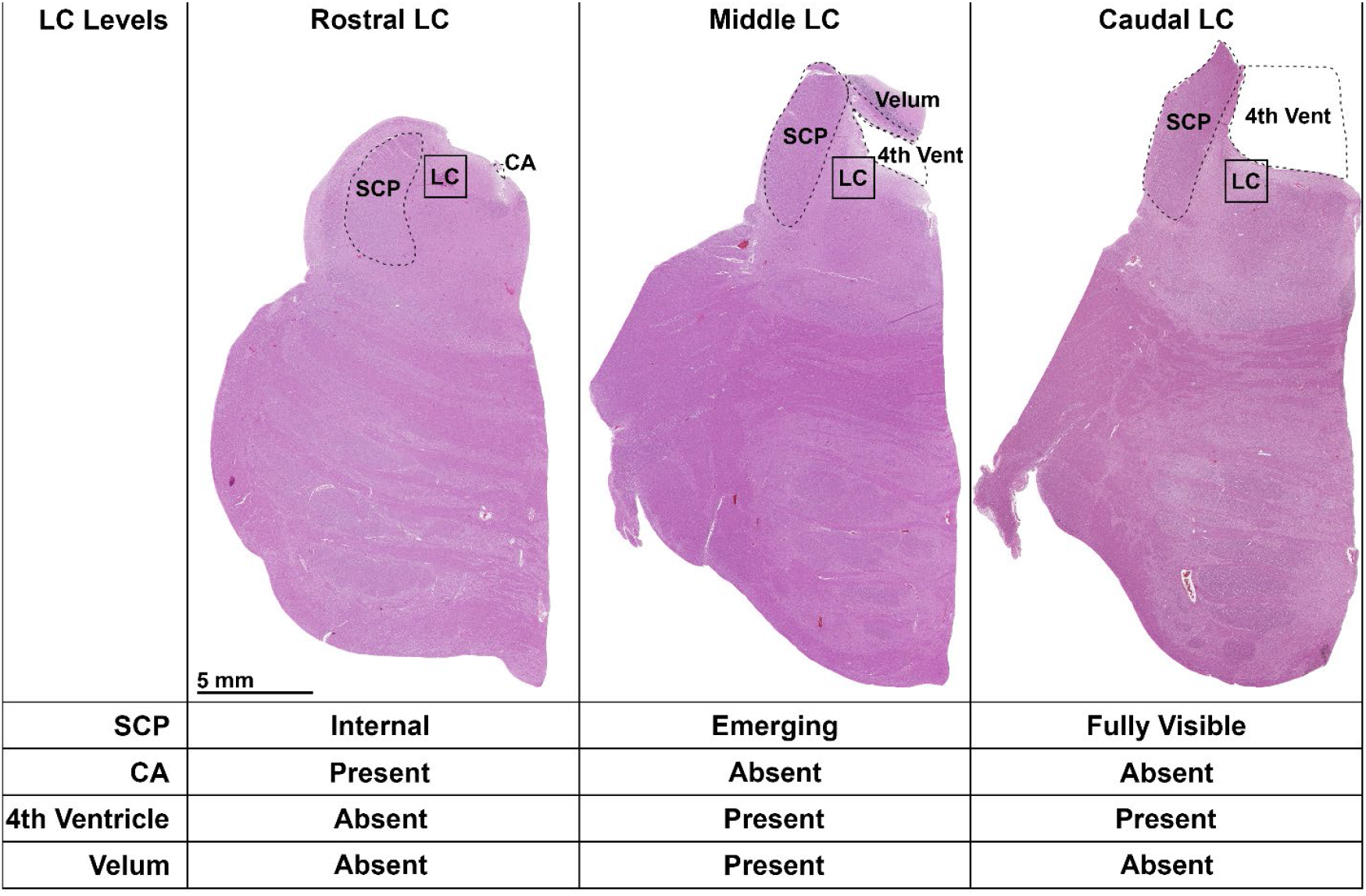
The rostral-to-caudal neuroanatomic extent of the locus coeruleus. The rostral locus coeruleus (LC) was neuroanatomically assigned based upon constrained superior cerebellar peduncle (SCP), presence of cerebral aqueduct and the absence of the 4^th^ ventricle and the velum. The middle LC was assigned based upon an SCP that was observed to emerge caudally, as well as the presence of the 4^th^ ventricle and the velum. The caudal LC was assigned when the SCP fully emerged caudally, presence of the 4^th^ ventricle, and absence of the velum. Neuroanatomic interpretation of LC levels utilized recommendations from German et al.[3] Acronyms: CA=cerebral aqueduct. LC= locus coeruleus. SCP=superior cerebellar peduncle.

**Figure S3.**
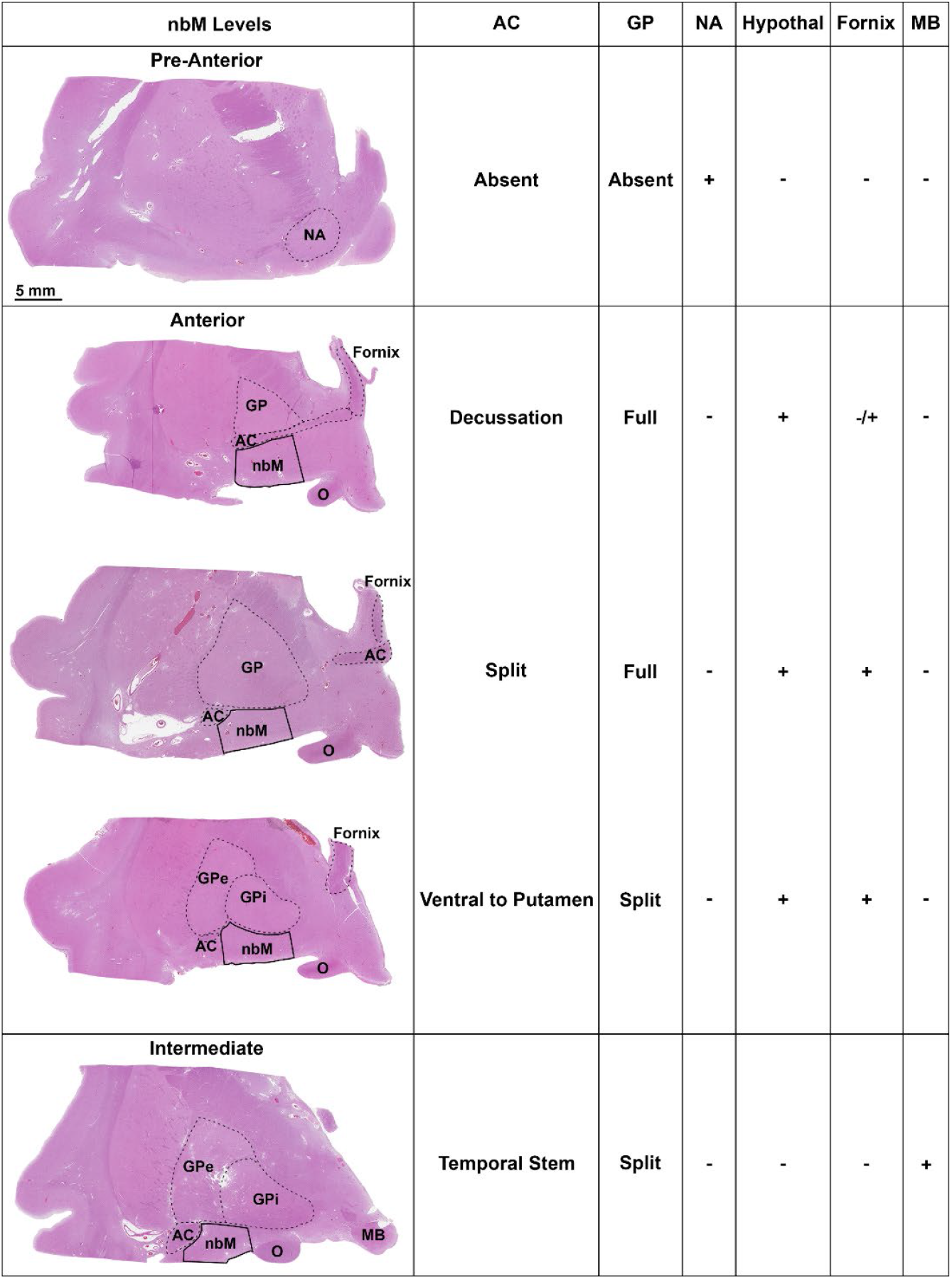
The anterior-to-posterior neuroanatomic extent of the nucleus basalis of Meynert. The pre-anterior nucleus basalis of Meynert (nbM) was neuroanatomically assigned based upon presence of the nucleus accumbens and the absence of both the anterior commissure (AC) and globus pallidus (GP). The anterior nbM was assigned based on an AC that was observed decussating, split, or ventral to putamen, a GP that was either full or split, and either the hypothalamus or fornix presence. The intermediate nbM was assigned based upon the AC observed descending into temporal stem, a split GP, and presence of the mamillary body present. Neuroanatomic interpretation of nbM levels utilized recommendations previously described in detail [5, 7]. Acronyms: AC=anterior commissure. GP=globus pallidus. Hypothal=hypothalamus. MB=mamillary body. NA=nucleus accumbens. nbM=nucleus basalis of Meynert. O=optic.

**Figure S4.**
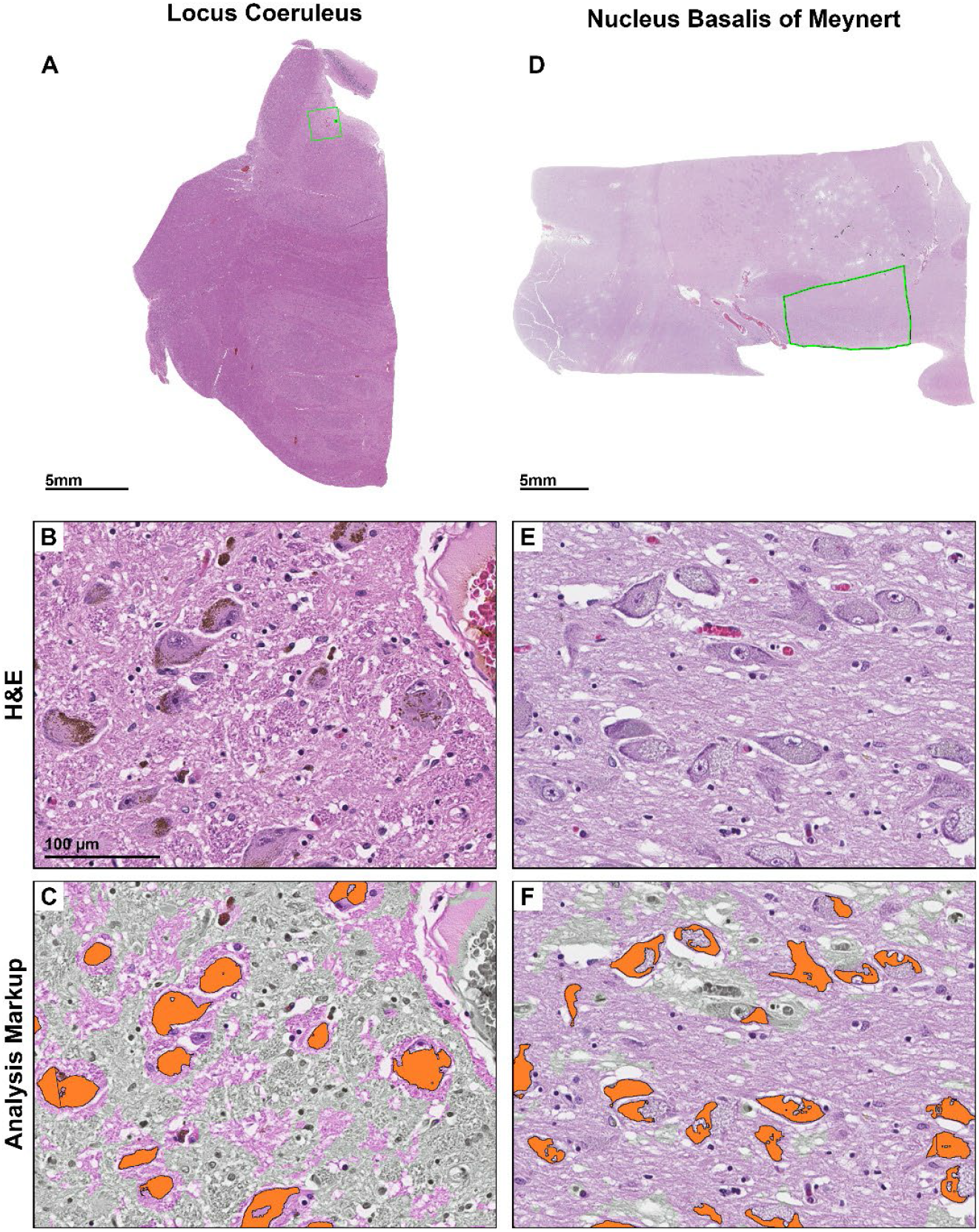
Digital pathology analysis of locus coeruleus and nucleus basalis of Meynert. Low magnification image of H&E-stained pons tissue section with 1800×1800 µm^2^ ROI in green **(A)**. A high magnification of the LC showing neuromelanin containing neurons **(B)**. The custom-designed neuronal count markup displays counted neurons in orange. Regions in gray were recognized to be neither LC neurons or neuropil by the pattern recognition software and were thus excluded from the neuronal count macro to enhance accuracy **(C)**. Low magnification image of the H&E-stained nbM tissue section with manually annotated ROI in green **(D)**. A high magnification of the nbM showing the neurons of interest. The neuronal count markup displays counted neurons in orange. Regions in gray were recognized by the pattern recognition software to be neither nbM neurons or neuropil and were thus excluded from the neuronal count macro **(F)**. Scale bar represents 5 mm in 1x view **(A**,**D)** and 100 µm **(B**,**C**,**E**,**F)**. Acronyms: LC=locus coeruleus. H&E=hematoxylin and eosin. nbM=nucleus basalis of Meynert. ROI=region of interest.

**Figure S5.**
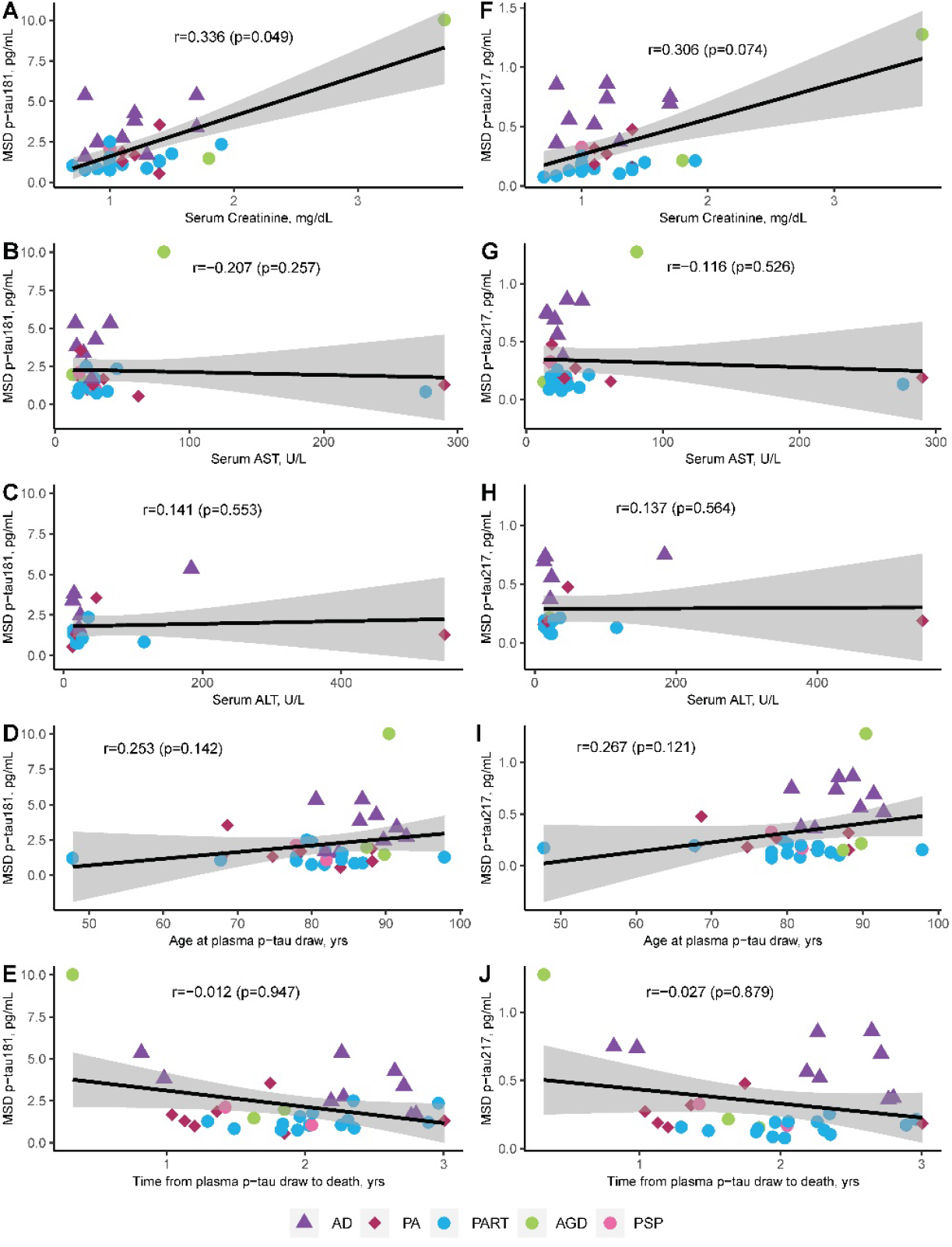
Evaluation of antemortem contributors to plasma p-tau variability. Plasma p-tau 181 **(A-E)** and plasma p-tau217 **(F-J)** was investigated for outliers contributing to variability as described in Supplemental Results (p 2). When examining serum creatinine levels from the kidney, we observed an outlier influencing the relationship with plasma p-tau levels. This outlier is displayed in subsequent graphs and included in Spearman correlations in this figure, but was removed from all subsequent analyses **(A, F)**. Levels of serum aspartate aminotransferase in the liver **(B, G)** and alanine aminotransferase in the liver **(C, H)** did not associate with plasma p-tau. Neither age at plasma p-tau draw **(D, I)** or time from plasma p-tau draw to death **(E, J)** associated with plasma p-tau levels. Spearman correlation and corresponding significance is shown. Trendline with 95% confidence interval was computed from a linear model. ALT=alanine aminotransferase. AST=aspartate aminotransferase. mg/dL=milligrams per deciliter. Acronyms: MSD=meso scale discovery. PA=pathological aging. PART=primary age-related tauopathy. pg/mL=picograms per milliliter. PSP=progressive supranuclear palsy. p-tau=phosphorylated tau for plasma levels. U/L= units per liter. yrs=years.

**Figure S6.**
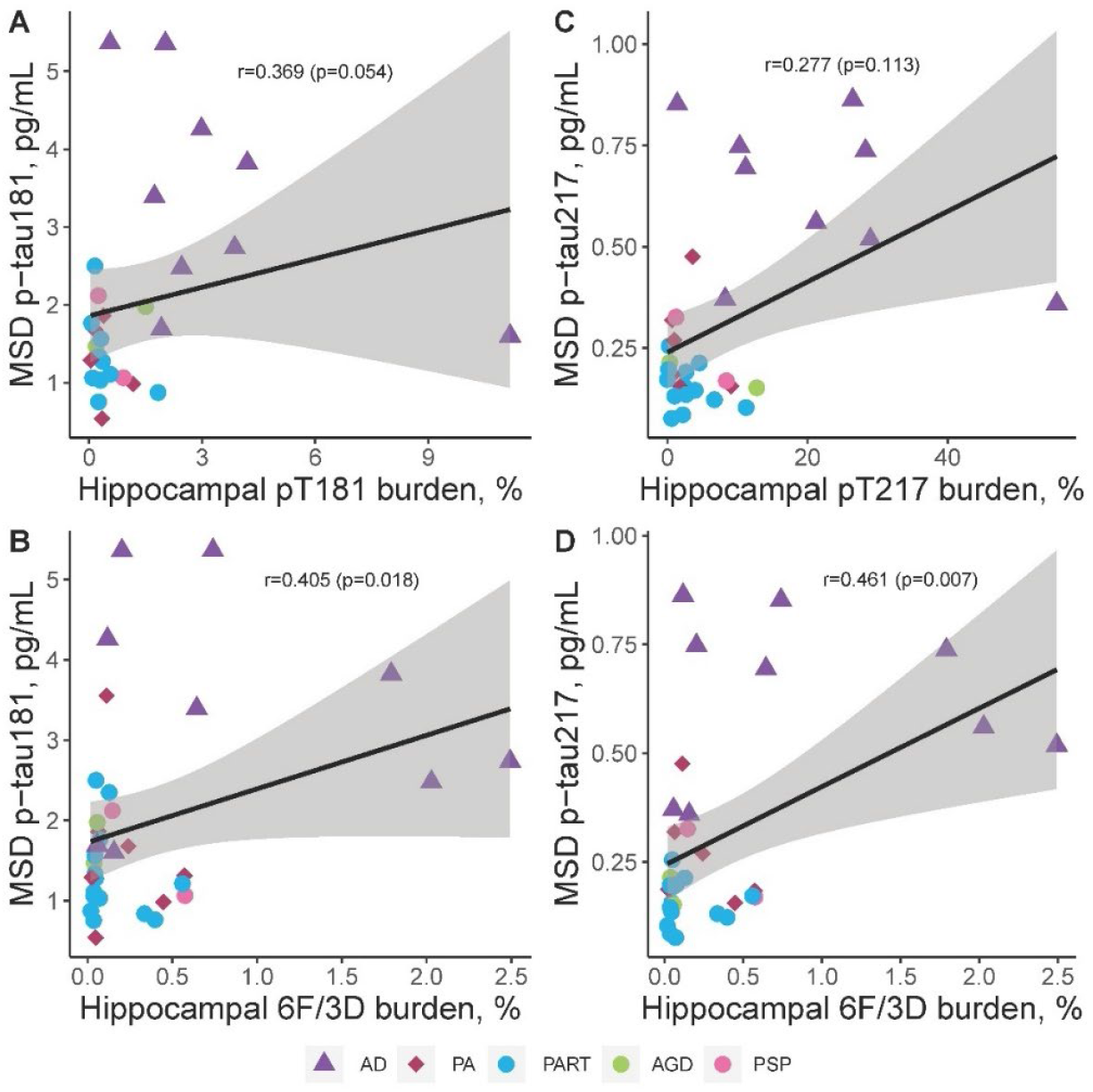
Neuropathologic evaluation of regional digital pathology measures of tau and amyloid-β pathology in comparison to plasma p-tau181 and p-tau217 in CA1-subiculum subsectors of hippocampus. Utilizing the same epitope to immunohistochemically evaluate regional tau pathology in hippocampus, tau burden measures were compared to p-tau plasma levels **(A, C)**. pT181 tau burden measures approached significance with p-tau181 plasma levels **(A)**, with association even not observed between pT217 and p-tau217 plasma levels **(C)**. Digital pathology measures of amyloid-β (6F/3D) were additionally compared to plasma p-tau levels **(B, D)**. Amyloid-β burden associated with ptau-181 **(B)**. The strongest overall association of digital pathology measures in hippocampus was observed between amyloid-β (6F/3D) and p-tau217 **(D)**. P-tau181 **(A-B)** and p-tau217 **(C-D)** were examined across all individuals studied with AD shown as triangles, PA as diamonds, and primary tauopathies as circles. Spearman correlation and corresponding significance is shown. Case with high creatinine was not included. Trendline with 95% confidence interval was computed from a linear model. Acronyms: AD=Alzheimer’s disease. AGD=argyrophilic grains disease. MSD=meso scale discovery. PA=pathological aging. PART=primary age-related tauopathy. PSP=progressive supranuclear palsy. pT=phosphorylated threonine for immunohistochemical measures of tau. p-Tau=phosphorylated tau for plasma levels.

**Figure S7.**
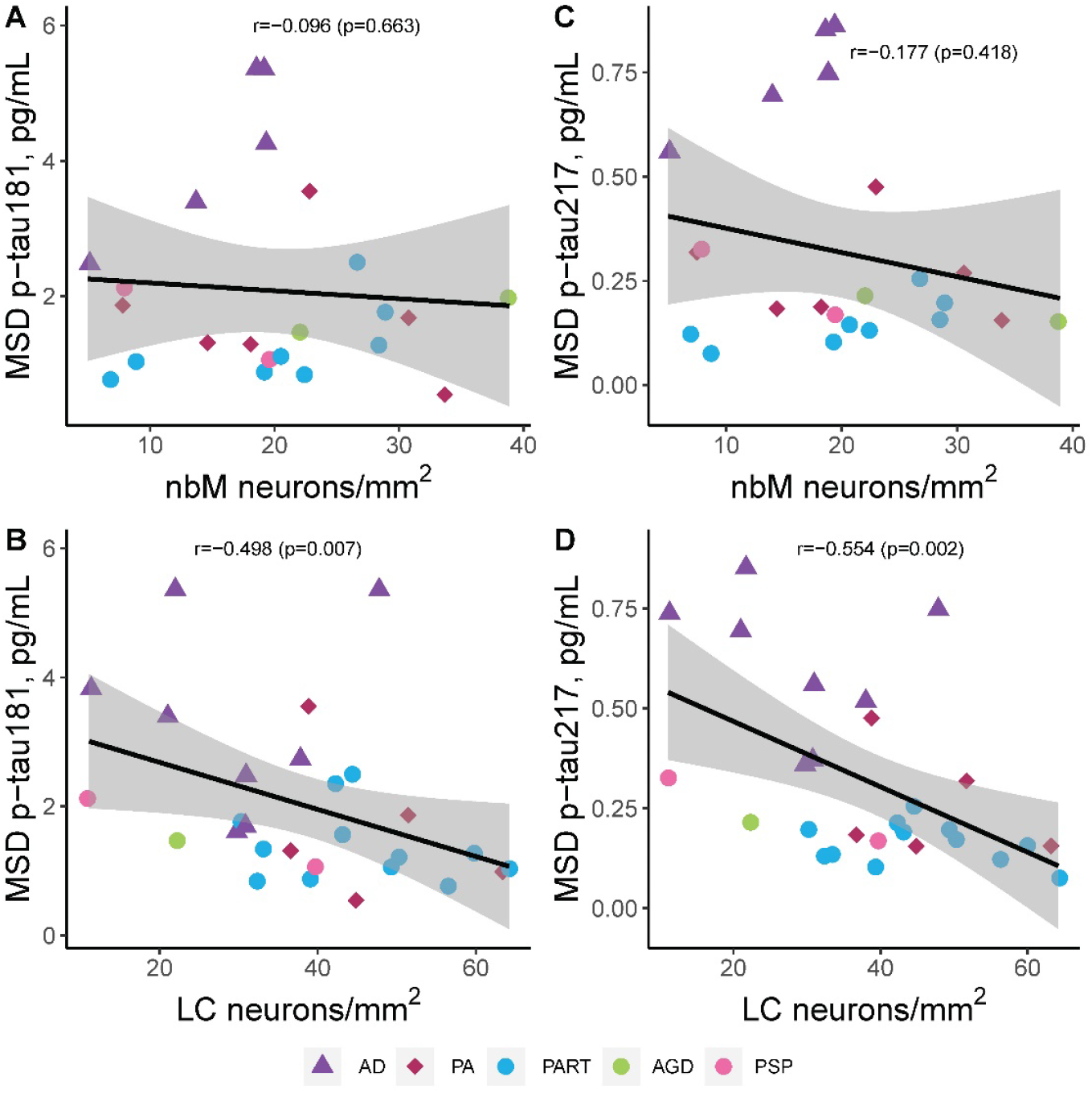
Neuropathologic evaluation of regional digital pathology measures of neurotransmitter hub neuron count in comparison to plasma p-tau181 and p-tau217 in nucleus basalis of Meynert and locus coeruleus. The nbM neuronal count did not associate with either p-tau181 plasma levels **(A)** or p-tau217 plasma levels **(C)**. Digital pathology measures of amyloid-β (6F/3D) were additionally compared to plasma p-tau levels **(B, D)**. However, lower LC neuronal count was observed to strongly associate with higher p-tau181 plasma levels **(B)** and higher p-tau217 plasma levels **(D)**. P-tau181 **(A-B)** and p-tau217 **(C-D)** were examined across all individuals studied with AD shown as triangles, PA as diamonds, and primary tauopathies as circles. Spearman correlation and corresponding significance is shown. Case with high creatinine was not included. Trendline with 95% confidence interval was computed from a linear model. Acronyms: AD=Alzheimer’s disease. AGD=argyrophilic grains disease. MSD=meso scale discovery. PA=pathological aging. PART=primary age-related tauopathy. PSP=progressive supranuclear palsy. pT=phosphorylated threonine for immunohistochemical measures of tau. p-Tau=phosphorylated tau for plasma levels.

**Figure S8.**
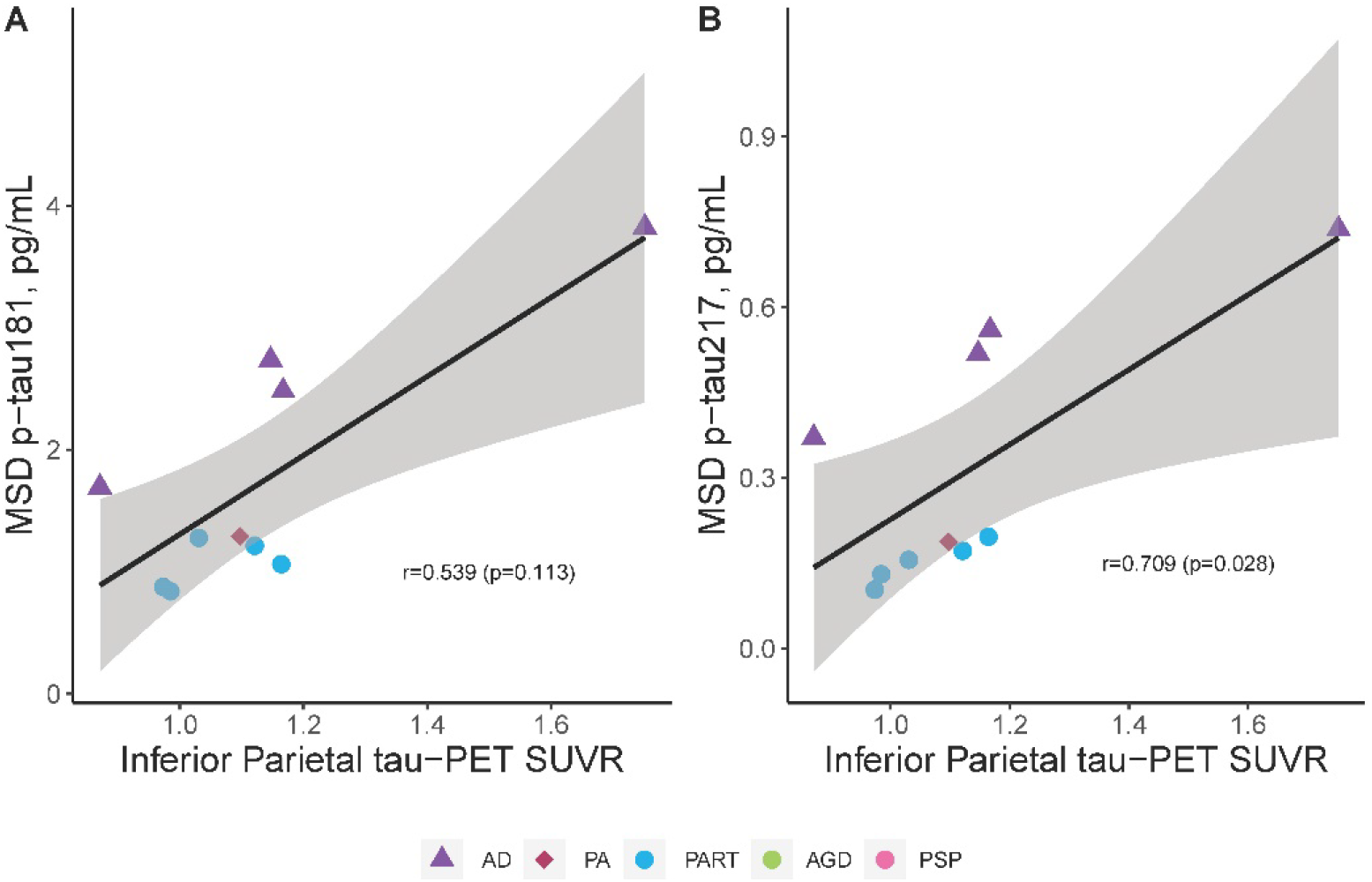
Evaluation of [^18^F]flortaucipir uptake in inferior parietal cortex and plasma p-tau levels. The relationship between regional [^18^F]flortaucipir uptake in inferior parietal cortex was investigated to extend the findings from immunohistochemical studies. We did not observe a relationship with plasma p-tau181 levels **(A)**, but did observe a strong relationship with plasma p-tau 217 **(B)**. Spearman correlation and corresponding significance is shown. Case with high creatinine was not included. Trendline with 95% confidence interval was computed from a linear model. Acronyms: AD=Alzheimer’s disease. AGD=argyrophilic grains disease. MSD=meso scale discovery. PA=pathological aging. PART=primary age-related tauopathy. PET=positron emission tomography. PSP=progressive supranuclear palsy. pT=phosphorylated threonine for immunohistochemical measures of tau. p-Tau=phosphorylated tau for plasma levels. SUVR=standard uptake value ratio.

**Figure S9.**
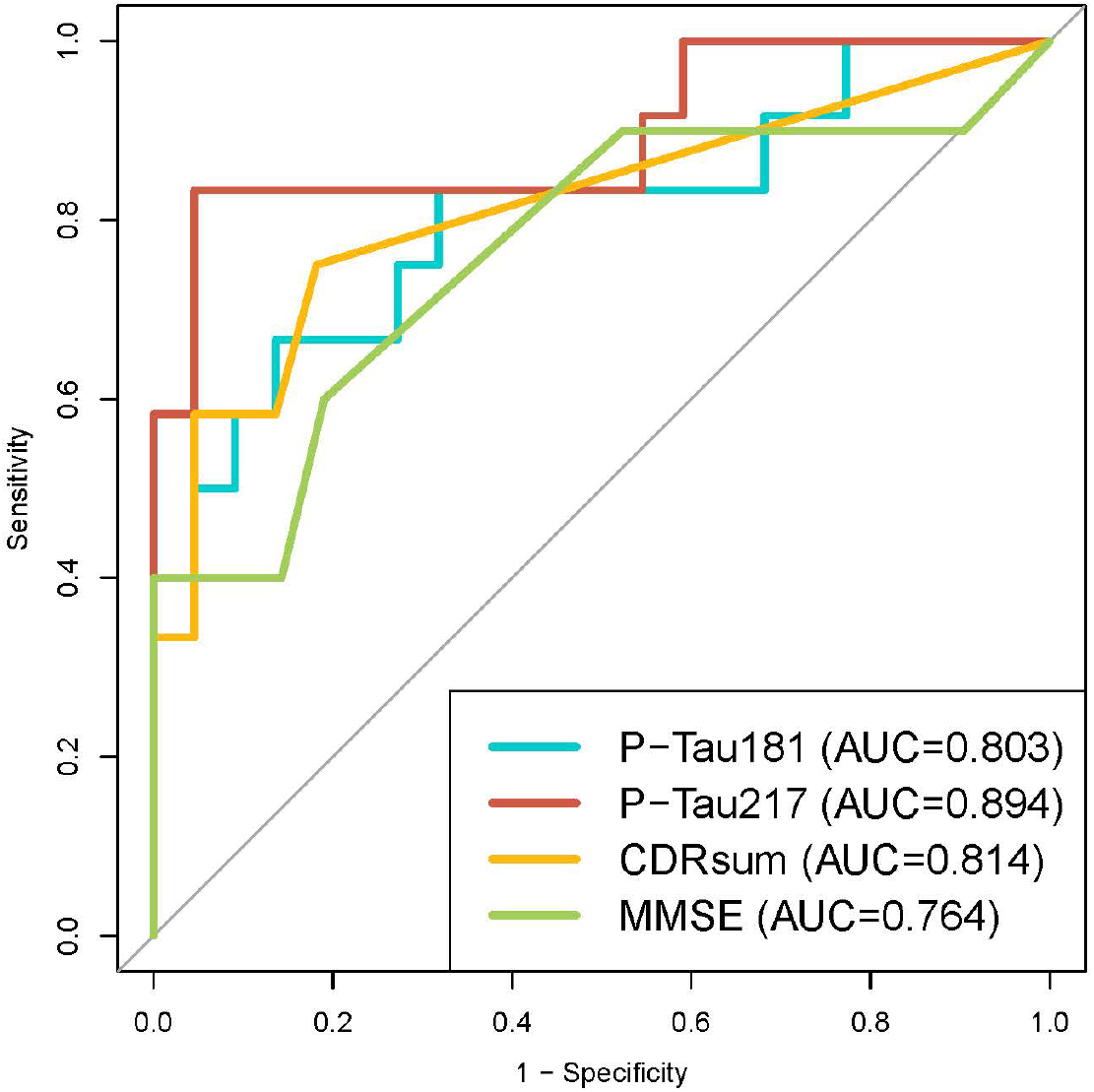
Predictive modeling of Alzheimer’s disease neuropathologic change. Predictive modeling of neuropathologic diagnosis of intermediate-to-high Alzheimer’s disease neuropathologic change compared to none-to-low show highest Area Under the Curve for p-tau217, followed by clinical dementia rating sum of boxes, p-tau181, and mini mental state examination. Case with high creatinine was not included. Given the sample size and use of logistic regression, plasma p-tau levels and cognitive scores were modeled individually, and were covariates not included. Acronyms: AUC=Area Under the Curve. CDRsum=clinical dementia rating sum of boxes. MMSE=mini mental state examination.

